# Deep Brain Stimulation restores information processing in parkinsonian cortical networks

**DOI:** 10.1101/2024.08.25.24310748

**Authors:** Charlotte Piette, Sophie Ng Wing Tin, Astrid De Liège, Coralie Bloch-Queyrat, Bertrand Degos, Laurent Venance, Jonathan Touboul

## Abstract

Parkinson’s disease (PD) is a neurodegenerative disorder associated with alterations of neural activity and information processing primarily in the basal ganglia and cerebral cortex. Deep brain stimulation (DBS) of the subthalamic nucleus (STN-DBS) is the most effective therapy when patients experience levodopa-induced motor complications. A growing body of evidence points towards a cortical effect of STN-DBS, restoring key electrophysiological markers, such as excessive beta band oscillations, commonly observed in PD. However, the mechanisms of STN-DBS remain elusive. Here, we aim to better characterize the cortical substrates underlying STN-DBS-induced improvement in motor symptoms. We recorded electroencephalograms (EEG) from PD patients and found that, although apparent EEG features were not different with or without therapy, EEG signals could more accurately predict limb movements under STN-DBS. To understand the origins of this enhanced information transmission under STN-DBS in the human EEG data, we investigated the information capacity and dynamics of a variety of computational models of cortical networks. The extent of improvement in decoding accuracy of complex naturalistic inputs under STN-DBS depended on the synaptic parameters of the network as well as its excitability and synchronization levels. Additionally, decoding accuracy could be optimized by adjusting STN-DBS parameters. Altogether, this work draws a comprehensive link between known alterations in cortical activity and the degradation of information processing capacity, as well as its restoration under DBS. These results also offer new perspectives for optimizing STN-DBS parameters based on clinically accessible measures of cortical information processing capacity.

**Significance statement:** Parkinson’s disease, a neurodegenerative disorder associated with a variety of motor symptoms, is due to the progressive degeneration of dopaminergic neurons. Neuronal networks in turn display abnormal activity associated with high excitability and abnormal synchronization. Treatments based on the electrical stimulations of deep brain nuclei (DBS) provide major symptomatic improvement, but their mechanisms of action remain unknown. Here, using mathematical models of the corticalcircuits involved, we show that DBS restores neuronal ability to encode and transmit information. We further show that movements from human patients can be better predicted from brain signals under treatment. These new theory and metrics open the way to personalized and adaptive DBS allowing to personalize stimulation patterns to each patient.

## Introduction

Parkinson’s disease (PD) is a neurodegenerative disorder associated with motor symptoms, including bradykinesia, rigidity, and tremor (1). These motor symptoms result from the progressive loss of dopaminergic innervation in the basal ganglia and alterations of neuronal activity in cortico-basal ganglia-thalamic circuits. In particular, at the cortical level, changes in excitability (2–4) and inhibitory activity (5–8), excessive oscillations and elevated synchronization in the beta frequency band (9–10), together with structural changes (11), have been reported in PD patients and animal models (12–20), as reviewed in (4,21). Furthermore, functional alterations of information processing during movement execution have been described in PD animal models, as illustrated by less precisely timed cortical spiking activity (22,23) and decreased sensitivity to incoming inputs (22,24,25). Altogether, this body of experimental evidence suggests that the detrimental changes in cortical dynamics arising in PD blur motor-related information and lead to a decrease in signal-to-noise that propagates to downstream circuits.

Chronic high-frequency stimulation of deep brain structures (called Deep Brain Stimulation, DBS), targeting the subthalamic nucleus (STN-DBS) or internal globus pallidus in the basal ganglia, has been shown to provide an effective symptomatic treatment in PD (1,26,27). However, the mechanisms of action of STN-DBS remain unclear (28–31). Various hypotheses have been proposed to explain the therapeutic effect of STN-DBS. In particular, several lines of evidence point out that, in PD patients and rodent models of PD, STN-DBS efficacy could be cortically mediated (32,18,13). Moreover, STN-DBS was shown to curtail primary motor cortex (M1) hyperexcitability in PD patients (33,3,34) and in PD rodent models (12,13), and to dampen cortical beta oscillations and bursting activity patterns (35–37). As a possible mechanistic explanation, STN-DBS is thought to recruit cortical GABAergic interneurons, as indicated by the restored cortical inhibition in PD patients (38,39) and by the increased activity in somatostatin (SST)-expressing cells in rodents (13). Hence, by restoring the dynamics of cortical networks, we hypothesize that STN-DBS could improve their information processing capacity, accompanying the alleviation of motor symptoms. In line with this hypothesis, a recent theoretical work showed that high-frequency stimulation, mimicking STN-DBS, restores the physiological activity of a neuronal network, by curtailing highly synchronized activity, and reinstates its information processing capabilities (40).

Here, we aimed at exploring this hypothesis based on EEG recordings from PD patients under pharmacological and STN-DBS treatments, and based on computational models. While EEG features did not show statistical differences between the OFF and ON-therapy conditions, we found that STN-DBS improved the ability to predict the type of movement that patients were planning to execute. To better understand the origins of the improvement in information processing capacity under STN-DBS, we tested the impact of STN-DBS on information encoding in multiple spiking models of cortical networks exhibiting various population activity and dynamics. We found that STN-DBS could reliably improve decoding accuracy of complex naturalistic inputs across most model configurations, and that STN-DBS parameters could be optimized according to the pathological activity profile of the computational model. Finally, we observed that STN-DBS could act by decreasing the excitability of pyramidal neurons, which in turn could shift the synchronization level of the network. This phenomenon was particularly exacerbated in PD networks exhibiting high levels of synchrony. Hence, our work finely investigates links between known alterations in cortical activity and the degradation of information processing capacity, and also opens new perspectives for more finely optimizing STN-DBS parameters, by relying on clinically accessible measures of cortical information processing capacity.

## Results

### Improvement of movement decoding from human EEG under STN-DBS

We set out to test the theoretical hypothesis that cortical circuits in PD patients display reduced information processing capabilities that are restored under STN-DBS. We aimed to quantify cortical information non-invasively in humans, using EEG, and compare these measures between control individuals and PD patients ON or OFF-therapy. To this purpose, we recruited a cohort of twenty human subjects, composed of 10 control individuals and 10 PD patients implanted with STN-DBS electrode (*Material & Methods* and **Table S1**), and performed multi-channel EEG recordings combined with electromyograms during the spontaneous execution of three movements. The three movements belonged to the clinical MDS-UPDRS severity scale for PD (41): finger tapping (item 3.4), fist clenching (item 3.5) and hand pronation (item 3.6) (see *Material & Methods and* **Movie S1** and **S2**). Electromyograms of the specific muscles involved (musculus extensor indici, flexor digitorum superficialis and profundus, and pronator teres muscles, respectively) were recorded and used to align EEG recordings relative to the onset of movement, allowing the extraction of movement-related potentials (**Fig. 1A**). PD patients were tested in both OFF and ON-therapy conditions. In OFF-therapy, patients were tested with STN-DBS OFF and weaned for more than 12 hours from their usual pharmacological antiparkinsonian therapy, while in ON-therapy, both STN-DBS and pharmacological medication were reinstated. While PD patients ON-therapy and control subjects successfully executed all three movements, two patients OFF-therapy were not able to perform enough repetitions for all three movements (< 50 trials) and one additional patient was not able to execute the hand pronation movement. In one last patient, the EEG signal was too noisy. These patients were excluded from further analyses since the associated dataset would not contain enough data points to be analyzed accurately. For statistical purpose (n=7 PD patients, n=10 control subjects), we centered most of our analyses of the EEG signals during the execution of two movements – finger tapping and fist clenching.

**Fig. 1:**
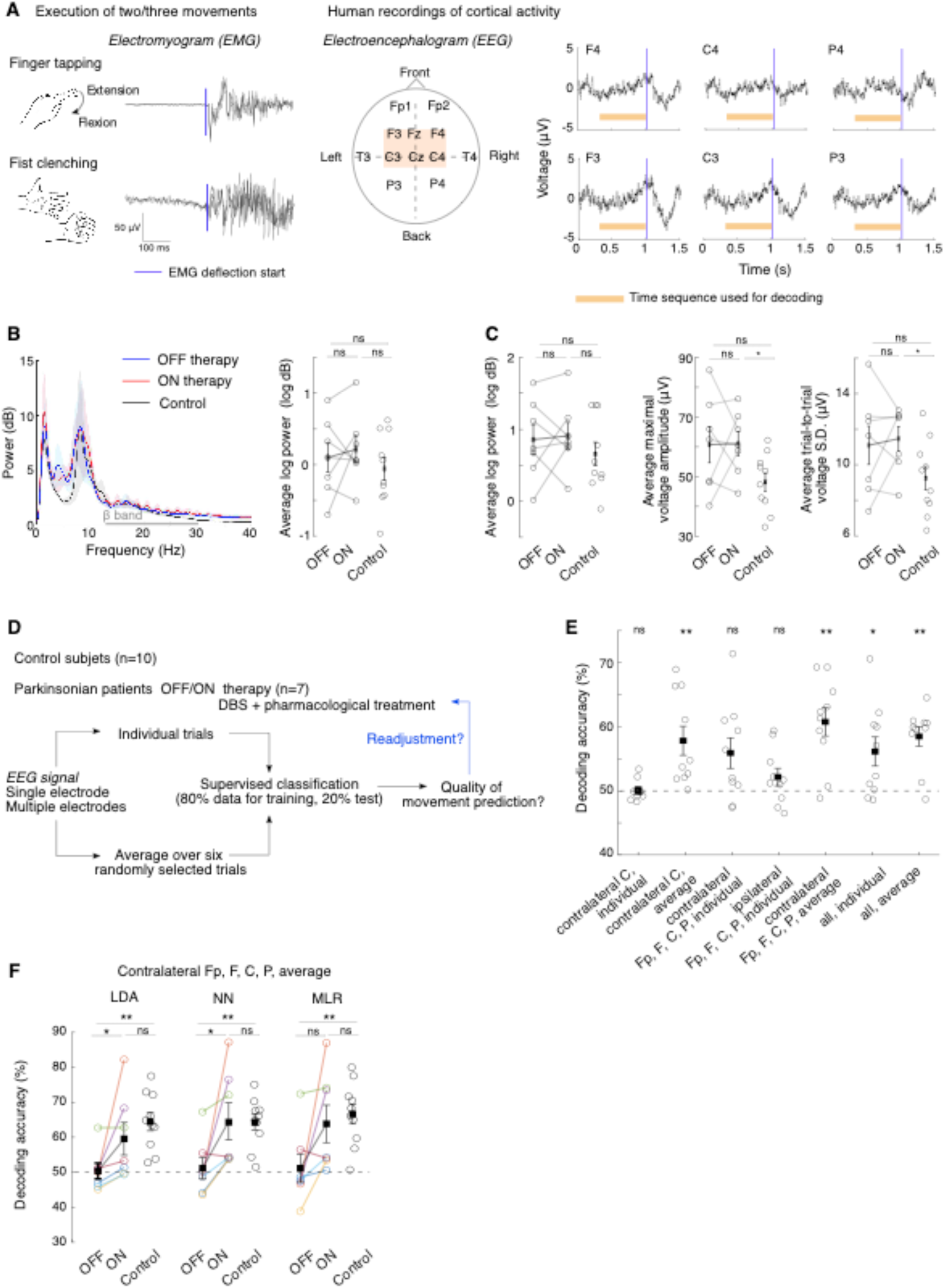
Altered information decoding from PD human EEG signals can be restored by STN-DBS and medication therapy. (**A**) Clinical protocol. Left: Illustrative averaged electromyogram (EMG) traces after alignment with the movement onset extracted from EMG deflections (blue line). Middle: Electroencephalogram (EEG) electrode positions (orange window: primary motor cortex). Right: Illustrative averaged EEG potentials aligned to movement onset (blue line). The time period used for classifying movements (initiation period) is indicated in orange. (**B**) EEG power spectrum from contralateral C electrode during the entire finger tapping session across conditions (left) and associated quantitative measure of mean beta-band power in the 13-30 Hz frequency range (right). Stats: ON/OFF therapy (paired Wilcoxon test, *p*=0.81), OFF/Control and ON/Control (independent Wilcoxon test, *p*=0.60 and *p*=0.60 respectively). (**C**) Quantitative measures of EEG features from contralateral C electrode during the initiation period for finger tapping movement. Left: Average beta-band power measured for each trial (ON/OFF: *p*=0.81; OFF/Control: *p*=0.37; ON/Control: *p*=0.42); Middle: Average maximal voltage amplitude measured for each trial (ON/OFF: *p*=0.94; OFF/Control: *p*=0.11; ON/Control: *p*=0.033); Right: trial-averaged standard deviation of the voltage (ON/OFF: *p*=0.94; OFF/Control: *p*=0.23; ON/Control: *p*=0.043). (**D**) Procedure for classifying movement identity from EEG signals. Signals either from a single electrode or from a combination of multiple electrodes, either from individual trials or from averaging of six randomly chosen trials, are used to train classifiers. (**E**) Decoding accuracy for control subjects (each dot corresponds to one subject) using linear discriminant analysis (LDA) for different input signals. One-sample Wilcoxon test (chance level: 50%, **Table S2**). **(F**) Decoding accuracy across three classifiers (LDA: linear discriminant analysis; NN: nearest-centroid and MLR: multinomial logistic regression) between patients ON and OFF therapy (paired Wilcoxon test) and with control subjects (independent Wilcoxon test). See **Table S3** for detailed statistics. Four electrodes (Fp, F, C and P electrodes) contralateral to the movement side were used.

To investigate whether EEG features varied across conditions, we quantified the average beta band power during each movement, either across the full session (**Fig. 1B** for finger tapping) or across individual trials, restricting the time window of the EEG signals to the pre-movement preparatory period (**Fig. 1C** for finger tapping and **Fig. S1B** for first clenching). A 700 ms-window preceding the EMG-detected onset of muscle activation was chosen in accordance with the start time of pre-movement readiness potentials reported in previous works (42). We focused on the electrode positioned over the primary motor cortex and contralateral to the movement side (labelled C electrode). During finger tapping, no differences in the average beta band power were observed across conditions (paired Wilcoxon test: OFF/ON: *p*=0.81 and *p*=0.81 for full session and individual trials, respectively; Wilcoxon test: OFF/Control: *p*=0.60 and *p*=0.37 respectively; ON/Control: *p*=0.60 and *p*=0.42); in particular no excessive beta-band power could be detected in PD patients OFF therapy. Alternatives measures of beta band power yielded a similar conclusion (**Fig. S1A**). In addition, we quantified the maximal amplitude of the preparatory movement-related potentials and did not find significant differences between OFF/ON-therapy conditions (*p*=0.94). Similarly, the trial-to-trial temporal variability, quantified by the standard deviation of the voltage signals across trials, did not reveal any modulation by STN-DBS (*p*=0.94). These observations were valid during fist clenching (**Fig. S1B**) and were consistent across electrodes (**Fig. S1C**).

Hence, although turning STN-DBS ON elicited an improvement in movement execution, no differences in basic EEG features were visible between the OFF and ON-therapy conditions. These observations are in line with previous EEG studies (43,44) (for review, see 43). We thus reasoned that quantifying cortical information might be more sensitive to capture latent signatures of movement planning in the EEG signals, and reveal differences in motor encoding between OFF/ON-therapy conditions. To this purpose, we compared the quality of decoding the identity of the executed movement from the EEG signals across conditions. Three supervised machine-learning algorithms – linear discriminant analysis (LDA), nearest centroid classifier (NN) and multinomial logistic regression (MLR) –, were trained and tested on EEG signals recorded during the preparatory period of the movement, to avoid biases due to the quality of movement execution (**Fig. 1D**). We ensured that movement identity could be decoded from control subjects and determined which configuration of input signals – either from individual trials or six-trial averages, and either from a single or a combination of electrodes – yielded the highest decoding accuracy (**Fig. 1E**, **Fig. S2A** and **Table S2**). When decoding two movements (finger tapping vs. fist clenching), we found that the signals from the electrodes contralateral to the movement were the most informative using LDA algorithm. Furthermore, combining the signals of four contralateral electrodes (Fp, F, C and P electrodes) yielded the best average accuracy across subjects (60.8%, *p* = 0.0059). Six-trial average allowed for better decoding accuracies than individual signals.

To test the impact of STN-DBS on cortical information processing, we compared the accuracy of decoding limb movement from EEG signals for each of the STN-DBS-implanted PD patients in both OFF/ON-therapy conditions. A significant improvement was found in the majority of patients (from 67 to 85%) once ON-therapy when decoding either two or three movements. More precisely, for 2 movements, out of 7 PD patients, 5 showed a significant increase in decoding accuracy using MLR and LDA algorithms, and 6 for NN; for 3 movements, out of 6 patients, 5 displayed increased accuracy for MLR and NN and 4 for LDA. On average, decoding of two movements performed by LDA and NN classifiers was significantly increased in the ON-therapy condition (LDA: 59.5% *vs.* 50.3 %, *p* = 0.0156; NN: 64.6 % *vs.* 51.1 %, *p* = 0.0312; MLR: 63.8 % *vs.* 51.1 %, *p* = 0.0781), and reached similar levels to those of control subjects (LDA: *p* = 0.193; NN: *p* = 0.962; MLR: *p* = 0.601). In contrast, decoding accuracy in the OFF-therapy condition relative to control was significantly worse (LDA: *p* < 0.001; NN: *p* = 0.0068; MLR: *p* = 0.0097) and did not differ on average to random chance level (**Fig. 1F**, **Fig. S2B** and **Table S3**). A similar trend was observed when considering other input configurations (individual vs. averaged trials, and using either all, or only the C electrode contralateral to the movement side) and when classifying the three movements. Yet, in these configurations, due to our small sample size, no statistical significance was observed between patients OFF and ON-therapy. Nonetheless, decoding accuracies OFF-therapy were significantly lower compared to control patients (**Fig. S2C and S2D** and **Table S3**). To verify whether these results could be affected by the quality of detection of movement onset, we repeated our analyses on control subjects by adding an artificial jitter of ± 50 to ± 200 ms to the original movement onset detected from the EMG recordings. While the decoding performance decreased, as expected from the addition of a source of randomness to the signal, accuracies remained consistently higher than that of PD patients OFF-therapy (**Fig. S2E** and **Table S4**), indicating that possible differences in the precision of detecting movement onset across conditions could not fully account for changes in decoding accuracy.

Overall, our EEG analyses indicate that hidden features, not easily captured by direct quantifications of movement-related potentials, may underlie the differences in decoding capacity observed between recordings of patients OFF vs. ON-therapy. These hidden features originate most likely from cortical dynamics, the access to which remains relatively limited in EEG.

### Emulating EEG results using motor cortex network spiking models

To emulate the results obtained from EEG data and further explore the links between cortical dynamics and information transmission, we developed a spiking network model of layer 5 of the primary motor cortex (M1), including three neuronal populations (**Fig. 2A**): pyramidal excitatory cells, parvalbumin (PV) and somatostatin (SST) inhibitory interneurons. The model relied on realistic parameters, based on our previous model (13) and published experimental data (**Tables S5-6**). Notably, consistent with experimental data, the model included the absence of a feedback connection from pyramidal cells to SST interneurons (45), as well as the fact that PV interneurons inhibit each other strongly while providing little inhibition to other interneurons, in contrast to SST interneurons (46,47). To recapitulate the diversity of cortical dynamics and activity regimes that are associated with different disease stages over the course of PD, different subtypes of PD as well as patient-to-patient variation in cortical wiring preceding PD onset, we generated multiple model configurations. We randomly varied three parameters of the original model, consistent with reports of altered excitation/inhibition balance in PD (2–8,12,13): the synaptic strengths reciprocally connecting pyramidal cells and PV interneurons (*w_e_* for pyramidal to PV and *w_i_* for PV to pyramidal cells), as well as the excitatory external input *I_ext_* received by pyramidal cells (**Fig. S3A**). This collection of network models generated a constrained repertoire of activity profiles (**Fig. 2B**), from highly regular periodic behaviors to asynchronous regimes, and from low to high firing rates. To organize the models according to the properties of the spontaneous dynamics they displayed, each network activity regime was characterized by (i) the mean firing rate of pyramidal neurons and (ii) the entropy of the average probability of pyramidal cells firing a spike, a proxy for synchrony (see also **Fig. S3** for additional quantifications and characterizations of the activity of the different networks). Two broad regimes emerged from this classification: oscillating regimes associated with low entropy (1-2 bits) and asynchronous regimes with high (> 3 bits) entropy. A small number of parameter configurations yielded relatively sparse activity (firing rate < 1 Hz) and intermediate entropy levels, creating a narrow tunnel connecting the two larger clusters of activity; no parameter combination generated intermediate levels of entropy with medium (1-4 Hz) to high (> 4 Hz) firing rates (**Fig. 2B**).

**Fig. 2.**
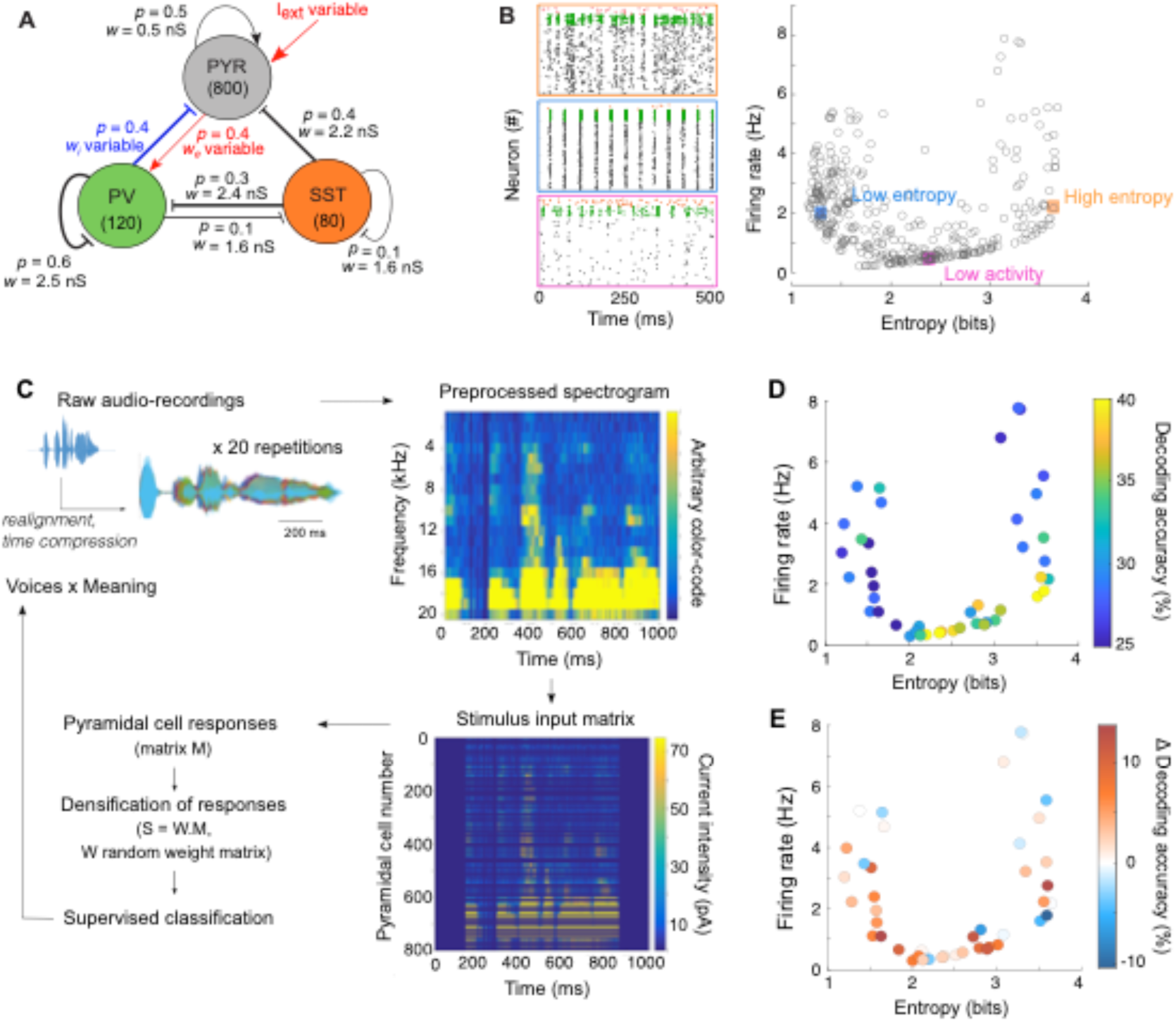
STN-DBS improves the decoding accuracy of naturalistic stimuli in a cortical spiking network. (**A**) Model architecture of layer V motor cortex, consisting of three cell types (PYR: pyramidal cells; PV: parvalbumin-expressing interneurons; SST: somatostatin-expressing interneurons) with population-specific connection probability *p* and synaptic weight *w* (arrow: excitatory, bar: inhibitory). Three parameters were varied: the external input to pyramidal cells *I_ext_*, the pyramidal-to-PV excitatory synaptic weight *w_e_*, and the PV-to-pyramidal inhibitory weight *w_i_*. (**B**) Left: Raster plots corresponding to distinct activity regimes: high entropy (and a medium firing rate) in orange; low entropy (with a similar firing rate) in blue, and low activity (with a low firing rate) in pink. Right: Scatter plot of the 2D network profile (across 370 network configurations) as a function of the mean firing rate and entropy of pyramidal neurons (the entropy was computed based on the average probability of pyramidal cells firing a spike within 5 ms time binned intervals, normalized to the average firing rate). (**C**) To emulate naturalistic stimuli, we used audio recordings converted into spectrograms and turned into an input to pyramidal neurons. This resulted in complex spatio-temporal patterns with trial-to-trial variability and latent variables (meaning of the sentence or voice of the speaker). (**D**) Decoding accuracy (color-coded) when trained to recognize meaning (4 meanings, chance level: 25%) from pyramidal cells activity in the OFF STN-DBS condition, as a function of the entropy and firing rate of the pyramidal cells OFF STN-DBS. Multinomial logistic regression was used as the supervised machine-learning classifier. (**E**) Changes in decoding accuracy under STN-DBS (ON – OFF) when trained on each dataset. Fixed STN-DBS parameters (200 pA, 130 Hz) were used.

Based on this broad benchmark of network profiles, we investigated the effect of STN-DBS on information processing in a subset of networks (n=44), by examining the network responses to complex and naturalistic stimuli, *i.e.* containing some periodicity and noise, encoding possibly multiple latent features and showing trial-to-trial variability. To this purpose, we built a set of complex non-stationary signals from audio-recordings (**Fig. 2C** and *Material & Methods*), that served as external inputs to pyramidal cells. The audio nature of the signals is indifferent to our purposes, but it captures features of naturalistic inputs, that could mimic the nature of inputs received by pyramidal cells of the motor cortex during movement preparation. Indeed, it provides us with a set of non-stationary signals (different spoken words) with trial-to-trial variability in the repetitions of the same sentences by the same individual, and two latent variables: the identity of the person speaking (the voice), and the meaning of the sentence. The voice or the meaning of the sentences, shared across different stimuli, can participate in creating common underlying features, such as the preferential activation of subsets of pyramidal cells (due to over-representation of a frequency for a given voice across sentences) or a common temporal pattern of activation (for a similar sentence across voices). We thus aimed at decoding these latent features by training the same supervised machine learning classifiers used for decoding movement identity from EEG signals (**Fig. 2D** and **S4A**, respectively). In absence of STN-DBS, the highest decoding accuracies (> 35%) were observed for very low-activity regimes (< 1 Hz), as well as high-entropy regimes with low firing rates (∼ 1-2 Hz). In contrast, the lowest accuracy scores were obtained for low-entropy (1-2 bits) or high-activity regimes (> 4 Hz), very close to random chance level (equal to 25% decoding accuracy).

Then, to mimic STN-DBS, we applied high-frequency pulse-like inputs to all three populations (13). Indeed, the currents associated with STN-DBS generates sequences of depolarizations along axons and synaptically-mediated depolarizations in the cortex, that originate from different anatomical routes: pyramidal cells can be antidromically activated through the hyperdirect pathway and orthodromically through the rapid STN-cortex pathway and the basal ganglia-thalamo-cortical route (48,49,12). Antidromic spikes can also excite deep-layer parvalbumin (PV) interneurons through the axon collaterals of pyramidal cells (antidromic axonal reflex) (50). SST interneurons have been shown to be activated under STN-DBS (13); while the mechanisms mediating the recruitment of SST interneurons remain to be investigated and most likely involve superficial layers, not included in our model, a high-frequency external current was added to SST interneurons to replicate experimental observations. For a fixed set of STN-DBS parameters (200 pA pulses at 130 Hz), decoding accuracy was increased under STN-DBS in 59% or 68% of networks when decoding the meaning or the voice respectively (**Fig. 2E and S4A**).

Altogether, we recapitulated the experimental observations made on EEG recordings by applying the same quantification of information – the decoding accuracy – to spiking network models in response to naturalistic stimuli. We found that STN-DBS consistently improved information transmission. These results allow us to further examine theoretically how the efficacy of STN-DBS can be optimized and predicted according to the properties of the network.

### Optimizing STN-DBS parameters for maximal decoding accuracy

In **Fig. 2E**, the extent of STN-DBS-induced change in decoding accuracy scores varied across network configurations. This variability may arise from differences in network sensitivity to external stimulation, which could be compensated by varying stimulation intensity. To test whether decoding accuracy could be maximized by varying STN-DBS parameters and explore how these optimal parameters relate to each network configuration, we varied both the amplitude and frequency of STN-DBS pulses (**Fig. 3A** and **Fig. S4B**) and evaluated changes in decoding accuracy in 20 network configurations exhibiting various levels of excitability and synchronization. The subset of network configurations examined here covered most regimes of the 2D-map presented in **Fig. 2B**. With optimal STN-DBS parameters, turning the stimulation ON always improved the decoding accuracy, with an average 60% (range from 30% up to 100%) increase in decoding accuracy compared to the OFF-DBS condition. We found that a single parameter – the total current of the stimulation (frequency multiplied by amplitude multiplied by pulse duration) – summarized well the effects of varying both STN-DBS frequency and amplitude: indeed, a notable peak in decoding accuracy arose for all network configurations when projecting STN-DBS amplitude and frequency onto the total current of the stimulation (**Fig. S4B**).

**Fig. 3.**
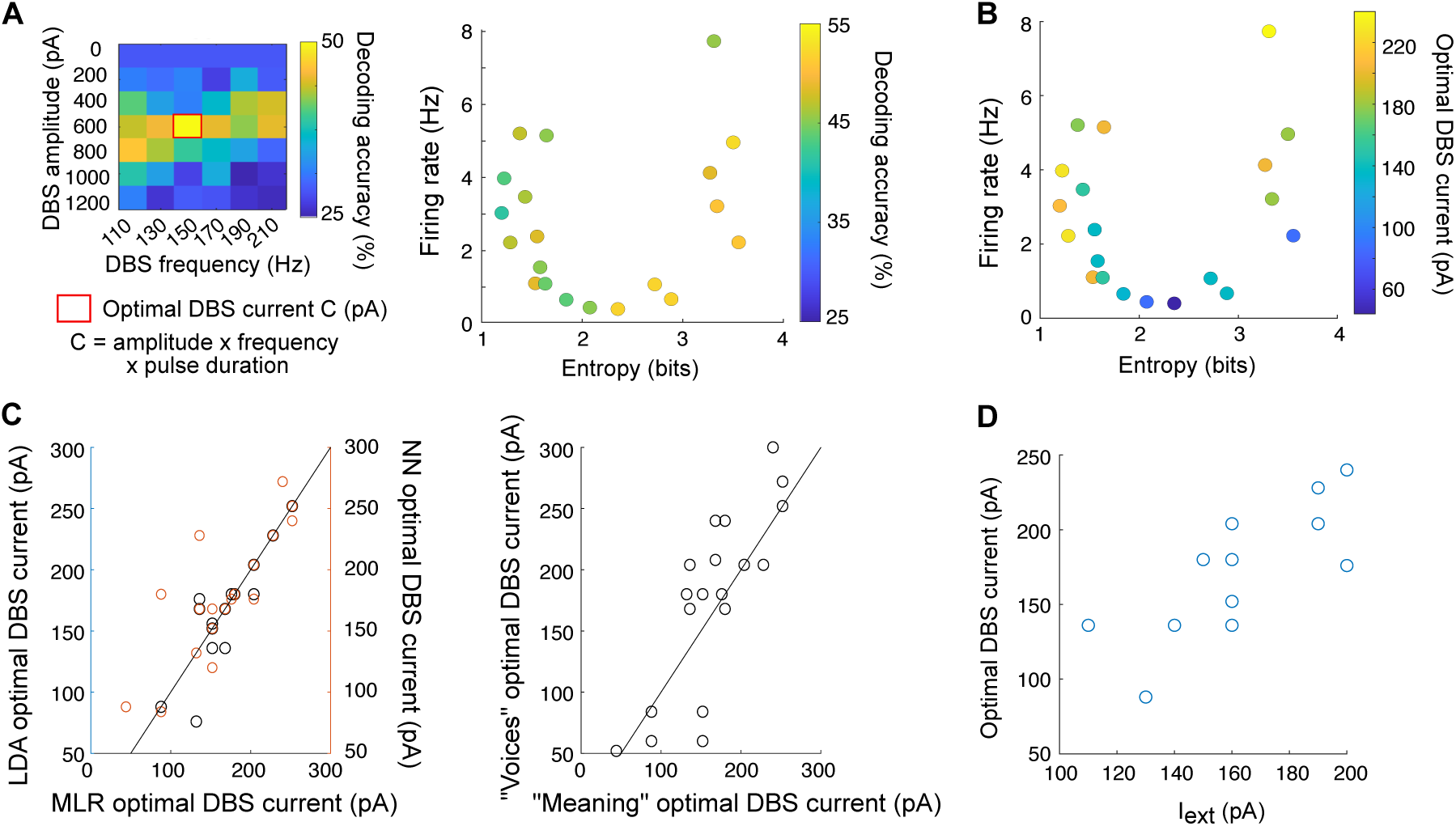
Optimizing STN-DBS parameters based on decoding accuracy and network parameters. (**A**) Left: Illustrative map of decoding accuracy (color-coded) of the multinomial logistic regression when trained to recognize meaning as a function of STN-DBS frequency (x-axis) and amplitude (y-axis) for one network configuration. An optimal STN-DBS current (red rectangle) corresponds to the amplitude x frequency x pulse duration parameters associated with the highest decoding accuracy. Right: Decoding accuracy (color-coded) obtained for the optimal STN-DBS current as a function of the entropy and firing rate of pyramidal cells OFF STN-DBS. (**B**) Optimal STN-DBS current (color-coded) as a function of the entropy and firing rate of pyramidal cells OFF STN-DBS. (**C**) Right: Correlation between the optimal STN-DBS current found by all three classifiers (Pearson’s correlation coefficient r and *p*-value: r(MLR, LDA) = 0.92, *p* < 0.001; r(MLR,NN) = 0.81, *p* < 0.001). Left: Correlation between the optimal STN-DBS current found when training MLR on meaning recognition *vs*. voice recognition (Pearson’s correlation coefficient r = 0.81, *p* < 0.001). (**D**) Correlation between optimal STN-DBS current and external current Iext added onto pyramidal cells (Pearson’s correlation coefficient r = 0.81, p < 0.001).

In line with the necessity for individualized clinical adjustments of STN-DBS parameters, information decoding could not be optimized across all network configurations using the same unique set of STN-DBS parameters. Indeed, the optimal STN-DBS parameters varied between network configurations (**Fig. 3B** and **Fig. S4B**). Importantly, for each network, the optimal STN-DBS current was robust to the choice of classifiers (Pearson’s correlation coefficient r = 0.81, p < 0.001 between MLR and NN and r = 0.92, p < 0.001 between MLR and LDA classifiers) and to the decoding feature (voice or meaning; Pearson’s correlation coefficient r = 0.81, *p* < 0.001) (**Fig. 3C**). These results support the hypothesis that the optimal STN-DBS current mostly depends on the intrinsic features of the network rather than the decoding task. In the same idea, networks with initially low firing rates necessitated the lowest currents (50-100 pA), since higher currents (> 150 pA) caused a complete and detrimental silencing of pyramidal cells (**Fig. 3B**). Among the three network configuration parameters we varied, the external excitatory input *I_ext_* targeting pyramidal cells was strongly correlated with the intensity of the optimal STN-DBS current (Pearson’s correlation coefficient r = 0.81, p < 0.001) (**Fig. 3D** and **Fig. S4C**). In the majority of networks, we also found a large gap between the high intensity of efficient STN-DBS current compared with the average strength of the stimulus, which was two to four times smaller. This gap was not prejudicial to information processing, underlining the strong filtering capacity of the cortical network relative to its shared STN-DBS input.

These analyses indicate that decoding of naturalistic inputs can be efficiently optimized by fine-tuning STN-DBS total current. The optimal STN-DBS current depended on the network configuration, and could be predicted from the level of cortical excitability.

### Linking STN-DBS impact on decoding accuracy to cortical profiles

To further decipher how STN-DBS affects the properties of the networks, and yields an improvement in information transmission, we studied a wider set of networks (those generated in **Fig. 2B**). We first investigated how STN-DBS impacted the firing rate and synchronization levels of pyramidal cells depending on network synaptic connectivity profile and intrinsic activity. Three main observations emerged from the comparison of OFF/ON STN-DBS conditions, using a fixed set of STN-DBS parameters (200 pA pulses at 130 Hz) (**Fig. 4A** and **Fig. S5A**): (i) high-frequency STN-DBS decreased the firing rate of pyramidal cells in 88 % of the network configurations, with at least a 50% reduction in firing rate in 62% of all networks; (ii) STN-DBS partially curtailed the synchronous activity of low-entropy regimes, resulting in increased entropy and decreased firing rate; (iii) high-entropy regimes became less entropic, yet without generating a regularized activity pattern imposed by STN-DBS. The strongest effects of STN-DBS were obtained in regimes dominated by a strong inhibition from PV to pyramidal neurons (high *w_i_*), and a low *w_e_*/*w_i_* ratio (Pearson’s r(*w_e_*/*w_i_*, ΔFiring/Firing) = 0.69, p < 0.001 and r(*w_i,_* ΔFiring/Firing) = -0.71, p < 0.001) (**Fig. 4B** and **Fig. S5B**). Conversely, a high *w_e_*/*w_i_* ratio was associated with an increase or no net change in firing rate under STN-DBS, while a strong external input *I_ext_* rendered pyramidal cells more resistant to desynchronization (Pearson’s r(*I_ext_*, ΔEntropy/Entropy) = 0.68, p < 0.001). Finally, STN-DBS did not alter the anti-correlation between pyramidal and PV firing rate, but accrued SST activity, hence enhancing SST inhibition on pyramidal cells (**Fig. S5C**).

**Fig. 4.**
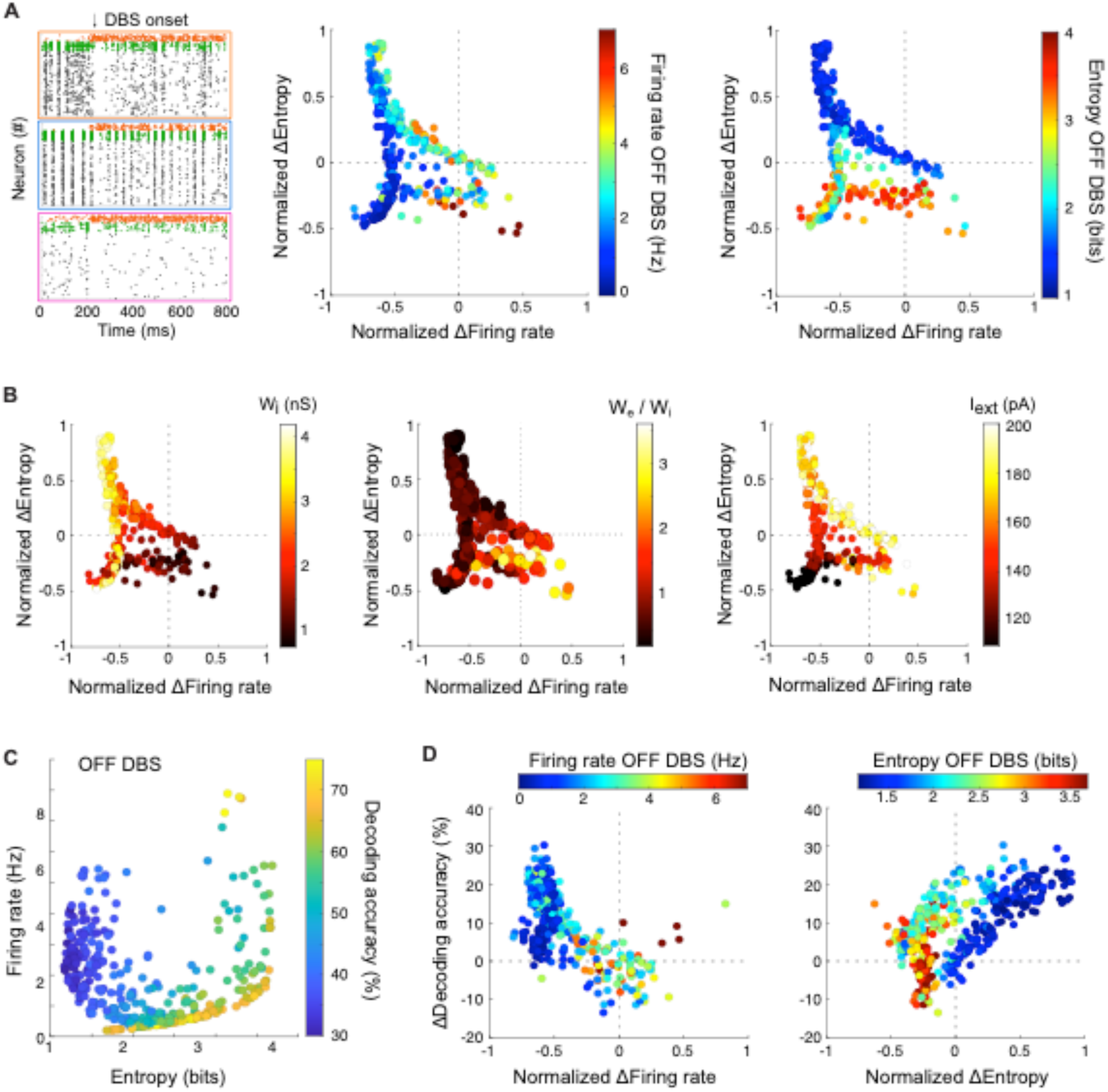
STN-DBS mitigates the pathological activity of cortical. networks by reducing firing rates and synchrony, thereby restoring information capacity. (**A**) Left: Raster plots of example networks before and after STN-DBS onset. The arrow indicates the time when STN-DBS input (200 pA, 130 Hz) is added to all neurons of the network. Normalized effects (ON-OFF/OFF) of STN-DBS on the firing rate of pyramidal cells (x-axis) and entropy (y-axis) of their responses depending on their initial regime (color-coded; middle: firing rate OFF STN-DBS; right: entropy OFF STN-DBS). (**B**) Impact of the parameters of the model (color-coded, left: *w_i_*, middle: *w_e_*, right: *I_ext._*) on STN-DBS efficiency for decreasing (x-axis) and desynchronizing (y-axis) the activity of pyramidal cells. (**C**) Supervised classification of simple stimuli using machine-learning algorithms on the responses of all pyramidal cells. Decoding accuracy in the OFF STN-DBS condition as a function of the firing rate (y-axis) and entropy (x-axis) of pyramidal cells OFF STN-DBS. (**D**) Changes in decoding accuracy under STN-DBS (ON – OFF) (y-axis) as a function of the normalized STN-DBS-induced change in firing rate (left) or entropy (right) (x-axis). Color-coded are the initial firing rate (left) and entropy (right) of pyramidal cells OFF STN-DBS.

Altogether, these observations suggest that STN-DBS is able to mitigate a wide spectrum of pathological cortical activity profiles. While STN-DBS was modeled as an external excitatory input to pyramidal, PV and SST neurons, its mechanism of action was paradoxically mediated by a decrease in excitatory tone. These results are in line with experimental findings, showing that the majority of pyramidal neurons decreased their firing rate whereas SST interneurons became activated under STN-DBS (13). We also found that the relative efficiency of STN-DBS depended on the intrinsic dynamics and connectivity of the network. Importantly, the impact of STN-DBS on cortical dynamics remained consistent when lowering the proportion of neurons receiving STN-DBS input (**Fig. S6A**).

To investigate how levels and patterns of cortical activity facilitate or hinder information encoding, we simplified the set of stimuli considered in the previous section and used constant or time-varying external stimuli as inputs targeting a subset of pyramidal cells (nine stimuli: two constant pulses, three Ornstein-Uhlenbeck processes and four deterministic ramps). We then compared the performance of supervised classifiers between OFF and ON STN-DBS. As observed in the decoding of naturalistic stimuli, high decoding accuracy (>55%, with random decoding accuracy equal to 11.1 %) was found in very low-activity regimes and in high-entropy regimes with low firing rates (∼ 0-2 Hz) in the OFF condition (**Fig. 4C** and **Fig. S7A**). In contrast, the lowest accuracy scores (< 40%) were consistently obtained for low-entropy regimes. STN-DBS efficiently improved the decoding accuracy of low-entropy regimes, with up to 160 % increase (**Fig. 4D** and **Fig. S7B**). In comparison to changes in excitability induced by STN-DBS, changes in entropy were a stronger predictor of changes in accuracy (linear regression model of ΔAccuracy based on MLR algorithm: ΔFiring: t = -1.02; *p* = 0.307; ΔEntropy: t = 9.58, *p* < 0.001). Indeed, both the sign and relative change in entropy governed the change in decoding accuracy. As such, in contrast to low-entropy regimes, networks characterized by a high entropy, slightly decreased upon STN-DBS application, were associated with a small decrease in decoding accuracy under STN-DBS. The robustness of these observations was supported by the high correlation between the decoding accuracy results obtained across all three machine-learning algorithms (**Fig. S7A**, Pearson’s r(MLR, LDA) = 0.96; r(MLR, NN) = 0.87; r(LDA, NN) = 0.86, *p* < 0.001). These results were also consistent when varying the proportions of neurons recruited by STN-DBS (**Fig. S6B**).

These analyses reveal that the low-entropy regimes, the most detrimental regimes for decoding external stimuli, are desynchronized by STN-DBS through a decrease in pyramidal cell activity, and constitute the regimes for which the impact of STN-DBS on information processing is the most prominent. Overall, the observations from our modeling framework, once adapted to the clinics, could have important implications for patient screening and STN-DBS parameter optimization.

## Discussion

PD has been characterized by functional alterations of information processing in the motor cortex during movement execution (22,23,25), together with changes in excitability and levels of synchronization (15,19,14,4,21). Based on previous experimental data (13), we further hypothesized that STN-DBS restores physiological activity patterns at the cortical level. To test this hypothesis and explore the links between information processing capacity and cortical activity in PD and under STN-DBS, we relied on EEG recordings from PD patients and data-driven simplified models of motor cortex.

While we did not find changes in basic EEG features, such as beta synchronization levels, across the OFF and ON-therapy conditions, in line with previous EEG reports (43,44), decoding movement identity from EEG signals, occurring before movement execution, was consistently more accurate in control and ON-therapy condition vs OFF-therapy. Indeed, in the latest condition, decoding accuracy reached chance level. These data thus indicated that cortical activity conveys more reliable information in control conditions and ON-therapy. Yet, such functional differences between conditions were not easily captured by basic signal statistics of the EEG, likely because of its limited spatial and cell-type resolution and low signal-to-noise ratio. This last observation prompted us to develop a modeling approach initiated in previously published works (13,40). To draw a close parallel to the EEG data, our metric for quantifying information processing in computational models relied on decoding known external stimuli targeting pyramidal cells based on pyramidal cells responses. In line with previous studies (16,51), we found that low-entropy regimes, characterized by highly synchronous activity across pyramidal neurons, were the most detrimental regimes for information encoding, whereas regimes of sparse spiking activity were associated to the highest decoding scores. Our results were consistent throughout the set of stimuli investigated, from simple deterministic currents to trial-to-trial variable and spatio-temporally structured inputs. Deficiencies in information processing can reflect various aspects of PD symptoms and can be related to altered network dynamics. Indeed, hyperexcitability or highly synchronized activity prevent external stimuli from affecting the spatio-temporal structure of the network responses to external inputs, making it more difficult to extract the relevant signals, as observed experimentally (22–24). Such abnormal activity associated to a decreased signal-to-noise ratio might cause an elevation in the detection threshold that would delay the initiation and slow down the sequential activation of voluntary movements*, i.e.* bradykinesia. In direct relation to decoding external inputs, PD patients often exhibit impairments in discrimination and perception of sensory stimuli, showing reduced sensitivity for detecting tactile and haptic stimuli as well as proprioceptive inputs, and reduced precision to differentiate between two stimuli (52–56). Altered decoding can be caused by a loss of specificity and of functional segregation of receptive fields, which in the motor domain can lead to the inaccurate recruitment of muscles and rigidity (14,57–59).

As observed from the increase in decoding accuracy, STN-DBS dramatically improved information processing in low-entropy regimes, and has milder effects in networks with similar activity levels but high-entropy. Interestingly, closely aligned to our information indicators, STN-DBS improves not only movement initiation and motor sequence execution, but also somatosensory and proprioceptive discrimination in PD patients (60,61). In line with our hypothesis, the STN-DBS-induced improvement in information processing was accompanied by a restoration of more physiological activity patterns: a decrease in the firing rate of pyramidal neurons was observed in most network configurations, mediated in part by the recruitment of SST interneurons, consistent with previous experimental work in rodents demonstrating the activation of cortical L5 SST interneurons and inhibition of pyramidal neurons under STN-DBS (12,13), and clinical studies (33,34,38,39). When applied to a low-entropy state, the decrease in firing rate further allowed a reduction in the level of synchronization of pyramidal cells spiking activity and the restoration of temporally richer dynamics, in accordance with experimental (18) and clinical findings (36,37).

We further investigated whether and how STN-DBS parameters could be adjusted to restore optimal information processing in cortical networks. We found that such optimization directly reflected the extent of STN-DBS-mediated desynchronization. In line with this result, a correlation between STN-DBS clinical efficacy and STN-DBS-mediated reductions in beta band oscillation power has been reported for STN (62,63) and at the cortical level (35). Our results also highlighted that STN-DBS efficiency is modulated by the initial regime of cortical activity. In particular, the higher the excitability of pyramidal cells (modeled by high values of the external current *I_ext_*), the higher the DBS current needed to be for optimized decoding. STN-DBS was also more effective at improving decoding accuracy in regimes characterized by a low entropy. Hence, we proposed that the levels of cortical excitability and synchronization could serve as predictive biomarkers of STN-DBS efficiency (64). Such features could be assessed in patients using transcranial magnetic stimulation and EEG recordings. If these predictions are first validated in the clinics, then statistics of cortical dynamics could be considered as additional criteria when screening PD patients for STN-DBS. We also observed that in our models the connectivity of the cortical circuit, controlling the excitation-inhibition balance, mattered in predicting STN-DBS efficiency: STN-DBS does not improve information encoding when pyramidal to PV connectivity is characterized by a low feedback inhibition and a strong excitatory drive. A tentative parallel can be made with known clinical and experimental observations: indeed, it is well acknowledged that patients who do not respond to L-DOPA are unlikely to benefit from STN-DBS (1). Despite the effects of L-DOPA in cortical circuits not being well elucidated, L-DOPA is known to affect cortical excitability and its oscillatory properties (65,66): D2 agonist increases PV-mediated GABAergic transmission onto pyramidal cells *in vitro* (67), while dopamine microinjections decrease the firing rate of pyramidal cells in anesthetized animals (68–70). Hence, in line with our modeling prediction, impaired or abnormally low GABAergic inhibition from PV interneurons might prevent L-DOPA efficiency and STN-DBS-mediated effects. In such regimes of activity, it may be interesting to test whether direct activation of neuronal subpopulations, for instance through the optogenetic activation of SST interneurons (13), might be more efficient than STN-DBS. Altogether, because our work explored a wide variety of network configurations and regimes of activity, we could better delineate and predict the range of STN-DBS efficiency, that could be tested in the clinics and used for patient screening.

Finally, our modeling framework opens the door to an alternative method for adaptive STN-DBS. Indeed, recent clinical research has relied on machine-learning algorithms and aimed at detecting the physiological states of vigilance of the patient (awake active, awake at rest, asleep) as well as the type of movement in combination with the level of beta bursts for adjusting STN-DBS parameters (71–73). We propose that the accuracy with which simple machine-learning algorithms decode movement identity from brain activity could also be used for adjusting STN-DBS parameters. This criterion might encompass a broader range of pathological markers compared to a single parameter (such as the amplitude of beta power or duration of beta bursts) currently used in adaptive DBS. Whether this method is best adapted to cortical signals (using EEG or electrocorticography, which has a better signal-to-noise ratio) or can be extended to other recording sites (for instance, STN) remains to be tested. Importantly, our results indicate that this method could be implemented non-invasively, in contrast to current procedures relying on LFP features and requiring higher signal-to-noise ratio measurements such as electrocorticography (72). The setup would allow to dynamically define optimal STN-DBS parameters based on cortical readability levels and an automatized estimation of movement improvement (74,75). Since optimizing stimulation parameters requires frequent visits to the hospital, a stressful environment for patients, especially in the first months after implantation, relying on an objective measure such as the accuracy of decoding movement-related information from brain signals should help both patients and neurologists.

Of course, implementing a model of cortical activity always relies on major assumptions and simplifications. Our model uses simplified equations for describing neural activity, and considers the impact of STN-DBS on cortex as an excitatory pulse-like current input delivered at high-frequency to all neurons. This simplification, together with the absence of local connectivity motifs among cell types, causes a homogeneity of responses within each population. Nonetheless, our model accounts for non-trivial network effects, such as the decrease in the activity of pyramidal neurons and PV neurons observed *in vivo* (13), and reproduced in a majority of cases *in silico*. In addition, our results remained robust when changing the proportion of neurons recruited under STN-DBS. Finally, we opted for a stimulus-based definition of *information*. If Shannon information theory has also been used in neuroscience, we chose a more functional quantification metric, which could be linked to different aspects of PD symptoms, as discussed above, and potentially be useful in the clinics. Interestingly, our previous work showed a high consistency between Shannon information and stimulus-response correlations (40).

Overall, our theoretical work highlights how a strong and extremely regular external input could nonetheless be compatible with the enrichment of intrinsic dynamics and information encoding of cortical networks, especially when applied to pathological cases. This effect is mediated through the filtering properties of reciprocal synaptic connections together with the recruitment of SST interneurons which efficiently reduces and redistributes pyramidal cells activity. In line with our previous works (40,13) and other computational models (76,77), STN-DBS mechanisms of action might be subtler than imposing an “information lesion” onto basal ganglia circuits (29). Beyond the theoretical analyses, our work could also have important consequences in the clinics, for patient screening and fine-tuning of STN-DBS parameters.

## Materials and Methods

### Spiking network models

The network model of the motor cortex L5 consists of 800 pyramidal, 120 PV and 80 SST cells. Neurons are modeled as adaptive exponential integrate-and-fire (78). The voltage of neuron *i* and its adaptation variable *w* satisfy the following system of differential equations:

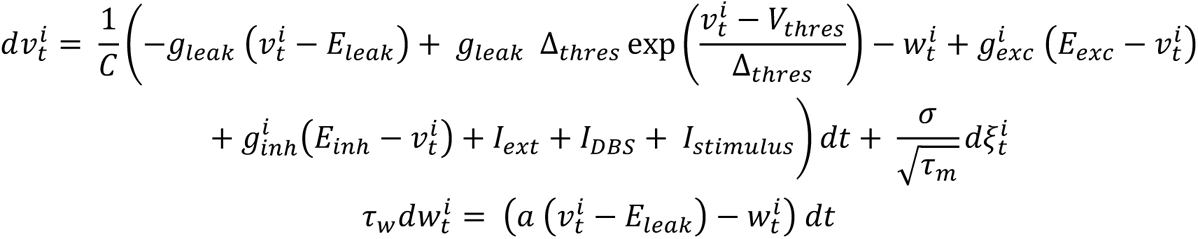

A spike is triggered when the voltage *v* reaches a threshold noted *V*_thres_. Upon firing, the voltage is reset to a fixed value noted *V_reset_*. Independent Gaussian white noise driven by a Brownian motion 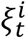 was added to the dynamics of each neuron to account for the variety of sources of fluctuations of the voltage (79). Choices of intrinsic parameters and connectivity parameters (**Tables S5** and **S6**) were guided by experimental data, as previously detailed in (13). The presence of a synaptic connection between two neurons was randomly drawn as a Bernoulli random variable with probability *p* to reflect the density of connections between various populations. The connection probability *p* ranged between 0.1 and 0.6. Synaptic weights were set to a fixed value *w*. Time constants for exponential decay of post-synaptic events were set to *τ_e_* = 5 ms and *τ_i_* = 8 ms for excitatory and inhibitory synapses respectively. To investigate different activity regimes, we randomly varied the connectivity weights between pyramidal cells and PV neurons (*w_e_*: synaptic weight from pyramidal cells to PV neurons; and *w_i_*: from PV to pyramidal cells) and the external input I_ext_ to pyramidal neurons.

To mimic the somatic impact of STN-DBS on cortical neurons, we added an external current *I*_DBS_ to the equation of the voltage variable in every cell of the network. More precisely, considering the periodic nature of STN-DBS-induced somatic currents, *I*_DBS_ corresponds to a series of square pulses (of 2 ms duration and 200 pA amplitude, repeated at 130 Hz, unless otherwise stated), corresponding to the depolarizations induced by each pulse of DBS. These depolarizations are the source of various anatomical and experimental observations of a cortical impact of DBS: first pyramidal cells can be antidromically activated through the hyperdirect pathway and orthodromically through the STN-cortex pathway (48, 49, 12). Antidromic spikes can also excite PV interneurons through the axon collaterals of pyramidal cells (50) and SST neurons are activated under STN-DBS (13). Finally, additional recruitment may come from a filtered STN-DBS input passing through the entire cortico-thalamic-basal ganglia loops. Due to these different putative activation pathways, *I*_DBS_ might impact neurons with specific delays, chosen to be: 0 ms for half of pyramidal and PV cells (fast antidromic pathways), 2 ms for the other half of pyramidal cells (orthodromic loops), and modulo 2 ms for SST interneurons (slow antidromic pathways to superficial layers and synaptic transmission). Note that the presence of such delays had no impact on network dynamics. Simulations of the network activity were done using a custom code developed in MATLAB R2022 (The Mathworks, Natick, MA, USA) using a Euler-Maruyama scheme with time step dt = 0.05 ms.

### Analyses

#### In the absence of a stimulus

For each network configuration, the firing rate and synchronization indices of the different neuronal populations in the absence of a stimulus were computed based on the average of 20 simulations lasting 300 ms (after an initial transient of 200 ms). Spike variance was estimated as the variance of the moving sum of all pyramidal cells’ spiking activity (with a window of 5 ms). This moving sum forms a time series typically characterized by peaks of activity, which are more or less exacerbated depending on the level of synchronized firing. Entropy was also calculated based on this moving sum, following Shannon’s definition, using 20 uniformly spaced bins from 0 to its maximal value. The Fano factor was defined as the average of the variance over the mean firing rate of all pyramidal cells over the variance for every 5 ms bin. The intrinsic frequency of the network was computed as the peak frequency of the modulus of the fast Fourier transform applied on the moving sum of pyramidal cells spiking. Unless specified otherwise, the firing rate and entropy measures are specifically applied to pyramidal cells only, and were used for a 2D description of neuronal activity.

#### Stimuli

In order to test the capacity of the network to discriminate and respond selectively to inputs, an additional current was injected to a subset of pyramidal cells (200 randomly chosen cells).

#### Simple stimuli

A set of nine simple stimuli were each presented during 500 ms: 2 constant inputs (70 and 60 pA amplitude), 4 ramps (from 0 to 70 and 0 to 60 pA upwards and downwards), and 3 Ornstein-Uhlenbeck noises (with different means, variances and time constants).

#### Naturalistic stimuli

Naturalistic stimuli originated from utterances recorded in MATLAB, obtained from 9 different voices. Overall, we could use 29 different sentences in French, with 4 shared meanings (“Je suis à Brandeis”, “J’habite à Paris”, “Je suis au Collège”, “J’enregistre ma voix”). Each sentence was repeated 20 times. Each audio recording lasted 2.5 seconds. To align all repetitions of the same sound stimuli, the beginning and end of each repetition were identified from a threshold crossing condition, common to all the repetitions of the same sound, except for some instances characterized by lower overall volume or higher noise, and for which thresholds were adjusted. For each aligned sentence, a spectrogram was computed based on the amplitude of the short-time Fourier transform of our signal: a windowing was made using segments of length 50, and 40 samples of overlap between adjoining segments. In total, 38 sampling points were used to calculate the discrete Fourier transform. The spectrogram was renormalized using a maximal thresholding value of 0.1 or 0.2 (this difference arises from the use of different audio-recorders). In the end, we obtained a spectrogram composed of 20 frequency bands, which was then fed as input to the network. Across stimuli, we decided to use the same set of frequency-dependent receptor cells, such that every 10 cells of the 200 receptor cells received the same frequency component over a 1000 ms of stimulus presentation. To make sure that all stimuli had similar input amplitudes (from 0 to about 70 pA) that were sufficient to elicit spiking activity, we multiplied the values of the spectrograms by 750 or 150. 2D maps as a function of DBS frequency and amplitude were obtained from the decoding of a subset of 12 stimuli (3 voices and 4 different meanings).

#### Information measures

We used the decoding accuracy of supervised learning algorithms trained on network responses to quantify the information capacity of the network. As described in (13), we used three supervised learning algorithms (nearest centroid classifier, multinomial logistic regression, and linear discriminant analysis) to estimate the efficiency with which neurons encoded various stimuli. Our approach consists of the following steps (**Fig. 2D**). First, our network responses are converted into a time-binned matrix M in which each row corresponding to a given pyramidal cell contains the number of spikes emitted for every 10 ms. The first 200 rows corresponded to the responses of the pyramidal cells directly activated by the stimulus. In order to reproduce the highly convergent cortical motor inputs received by striatal neurons, we densify the responses by contracting the M matrices of 800 × 50 (for simple stimuli) or 800 x 100 (for naturalistic stimuli) dimensions into a 100 × 50 (or 100 x 100) matrix S, defined as: S = W.M, where the weight matrix W is a random matrix, identical for all stimuli presentations, with each element generated from the uniform distribution on the interval [0, 1]. These S matrices are then used as inputs for the supervised-learning algorithms, (80).

For each condition, the dataset consisted of 60 repetitions for each of the 9 simple stimuli or 20 repetitions for the naturalistic stimuli (with the same network parameters and convergence matrix W, but independent realizations of the intrinsic noise ξ, and in the case of naturalistic stimuli trial-to-trial variable input patterns). The classifiers were trained to discriminate the population response given the stimulus on 80% of the data sample, using stratified k-fold cross validation (*k* = 5). This procedure was repeated using 5 independent seeds. In the case of naturalistic stimuli, the classifiers could be trained on different rules: recognizing voices, recognizing meaning, or recognizing both attributes. Training and testing of the classifiers were run using scikit-learn and keras packages in Python 3.5 (Python Software Foundation, www.python.org).

### EEG acquisition

#### Participants

Twenty human subjects participated in the study (see **Table S1** for detailed characteristics). All subjects gave informed consent for participating in the study. The study protocol was approved by the Ethics Committee of the APHP (Assistance Publique-Hôpitaux de Paris, Paris, France), commission CPP 09-2022. It is registered on ClinicalTrials.gov as NCT05284526. The cohort was composed of two groups:

- 10 control subjects
- 10 PD patients with implanted STN-DBS and dopaminergic treated, recorded on four sessions: OFF STN-DBS and OFF medication; ON STN-DBS and OFF medication treatment; OFF STN-DBS and OFF medication; ON STN-DBS and ON medication. In this study, we focused our analyses exclusively on the OFF STN-DBS and OFF medication condition (OFF therapy) and the ON-STN-DBS and ON medication condition (ON therapy).

Patients with no cognitive impairment were selected (Montreal Cognitive Assessment score > 24). PD patients were diagnosed according to the current diagnostic criteria (81,82) and were recruited from the Neurology Department of the Avicenne University Hospital.

Recordings were performed at the Service de Physiologie, Explorations Fonctionnelles et Médecine du Sport, Avicenne University Hospital. The motor function of all patients was assessed for each condition before performing the task, using the International Parkinson and Movement Disorders Society Unified Parkinson’s Disease Rating Scale part III (MDS-UPDRS III). During the experiment, patients were in a practically defined ‘OFF medication’ state, after overnight withdrawal (at least 12 hours) of PD medication. Sessions ‘ON medication’ started 45-60 minutes after resuming the usual treatment.

#### Post-hoc exclusion

Two PD patients were unable to perform any movements OFF therapy and were thus excluded from our analyses, since we could not reliably identify the time segments preceding movement. This increased uncertainty and the poorer performance resulting from it would have made our estimates of the PD patients decoding accuracy even lower, and it seemed a fairer comparison to estimate the accuracy only for patients able to perform the movements. In another PD patient, the SNR of the EMG recordings during the pronation of the hand movement was too low for detecting muscle activation in STN-DBS OFF condition, due to the inability of the patient to clearly execute the movement; this patient was not included when analyzing all three movements for the same reasons. Finally, in one PD patient, large noisy fluctuations in EEG signals due to poor grounding of the electrodes prevented the extraction of clean EEG traces.

#### Task

Subjects were positioned in a comfortable chair, half-seated with legs extended. They were asked to perform three different movements: index extension, clenching of the fist and pronation of the hand. The task was divided into eight blocks. Each block consisted of repeating each movement without external trigger, in a self-paced manner: when feasible, it was constituted of 25 repetitions of first index extension (musculus extensor indici muscle), then 25 repetitions of clenching of the fist (flexor digitorum superficialis and profundus muscles), and finally another 25 repetitions of the pronation of the hand (pronator teres muscle). These hand movements correspond respectively to the 3.4, 3.5, 3.6 items of the MDS-UPDRS III (41). Overall, we could obtain at best about 200-250 repetitions for each movement. Subjects were asked to close their eyes and relax during movement execution. See for an example **Movie S1** and **S2**.

#### EEG recordings

Signals were recording using a 15-channel EEG system (System PLUS Evolution manufactured by Micromed SpA, Version 1.07.00), sampled at 1024 Hz. Bipolar EMG of musculus extensor indici, flexor digitorum superficialis and profundus, and pronator teres muscles, on the same side of movement, were also recorded. EEG cup electrodes (Capsulex 333102, MEI) were positioned manually and fix with plasters to ensure good stability across the different recording sessions. The following sites were recorded: {‘Fp1’, ‘Fp2’, ‘F3’, ‘F4’, ‘Fz’, ‘T3’, ‘T4’, ‘C3’, ‘C4’, ‘Cz’, ‘P3’, ‘P4’}. A reference electrode was positioned on the skin over the mastoid bone.

#### Extraction of movement-related potentials

EEG signals were processed using custom MATLAB scripts. Signals were first band-pass filtered (1-100 Hz). 50 Hz line noise was removed when necessary (notch filter). Continuous EEG analysis resulted from the concatenation of movement execution periods, which contained no evident artefact (cut-off amplitude of 80 µV). Movement-evoked potentials were obtained after determining the onset of movement for each trial using EMG recordings. On most cases, the SNR was sufficient to perform an automatic analysis based on a custom MATLAB script (using in particular *findchangepts* function). In other cases, in particular when the EMG trace was constituted of multiple upward and downward phases for a single movement, a manual inspection on the EMG trace was necessary to detect the onset of movement. Each movement onset was then verified based on video recordings alignment.

#### Statistics of EEG signals

To compute beta band power, a fast Fourier transform was applied to the EEG voltage from individual electrodes, either on the full session of a given movement or on individual trials (using a 700 ms period preceding movement onset). Average beta band power was obtained by averaging the power in the 13-30 Hz frequency range (**Fig. 1B and C**), or in the 17-23 Hz range (**Fig. S1A**). As an alternative measure, the power of the highest peak in the 13-30 Hz band of the power spectrum was also extracted (**Fig. S1A**). In the 700 ms-long movement-related potentials of the preparatory period, the maximal voltage amplitude (the envelope between maximal and minimal voltage values) of each trial were calculated and then averaged to yield a single data point per patient and per movement (**Fig. 1C** and **Fig. S1B**). Finally, the standard deviation of voltage fluctuations across trials was averaged over the 700 ms window (**Fig. 1C** and **Fig. S1B**).

#### Decoding EEG signals

Movement-related potentials, either individual or averaged over six randomly chosen trials (average-based trials), constituted our input data stream for our decoding analyses. We selected the 700 ms period preceding movement onset, such as to focus exclusively on cortical preparatory activity. This way, signal decoding was not influenced by the quality of movement execution itself. In addition, these preparatory signals were not contaminated by low-frequency movement-related artefacts, which were clearly visible in some subjects. Signal decimation by a factor of 3 did not affect decoding accuracy and was therefore applied to speed up analyses. Signals used originated from individual electrodes (contralateral to the movement for Fp, F, C and P sites) or midline (Fz and Cz), a set of four contralateral or ipsilateral electrodes (Fp, F, C and P) or all electrodes (*i.e.* bilateral Fp, F, C, P, Fz and Cz electrodes, excluding T3 and T4 electrodes). In the case of decoding movement identity from EMG electrodes, the signal from the three EMG electrodes was used in a window of 700 ms (but compared to EEG traces focusing solely on the preparatory period, the start of the window used for EMG traces was 200 ms delayed so as to include the beginning of the movement). Jittered EEG data were obtained by realigning EEG traces to jittered movement onsets. The jitter was randomly defined across trials and drawn from a uniform distribution bounded at ± 50, 100 or 200 ms (from the initially detected movement onset). Decoding was then performed using either a fixed number of trials over all the population or a fixed number of trials per patient. Trials were then randomly assigned to the training (80%) or test (20%) data set, using a stratified k-fold cross-validation (k=5). This procedure was then repeated using 5 independent seeds. The training and testing of the classifiers were run using scikit-learn (https://scikit-learn.org/) and keras (https://keras.io/) packages in Python 3.5 (Python Software Foundation, www.python.org). 100 individual trials or 20 average-based trials were used for comparing, in control subjects, the decoding accuracy between electrode sites and configurations. For comparing patients OFF and ON therapy and control subjects, a patient-based selection was performed, after ensuring the absence of correlation between the number of training trials and decoding accuracy. The number of individual trials varied from 46 to 251 (and consequently average-based trials from 7 to 41). We ensured the absence of a significant correlation between decoding accuracy and the number of training templates (**Fig. S2B**). We used the same three algorithms previously described for decoding network spiking responses.

### Statistical Analysis

Results are expressed as mean±SEM. Statistical significance was assessed using R2022 (The Mathworks, Natick, MA, USA) or Jamovi (http://www.jamovi.org). For analysis of experimental data, paired or independent two-sampled two-tailed Wilcoxon sign rank were used to compare two distributions, or a one-sample Wilcoxon test for comparing decoding accuracy to chance level. Statistics of decoding results are summarized in **Tables S1**, **S2** and **S3**. Linear regression models were used to assess the dependency of one variable onto another (or multiple others). Correlations were assessed using Pearson or Spearman correlation coefficients.

## Data Availability

All data produced in the present study are available upon reasonable request to the authors

## Code availability

Custom MATLAB codes used for the computational model will be made publicly available on GitHub.

## Acknowledgments

We thank J.E. Rubin, P. Miller, the members of the LV and JT laboratory for their helpful suggestions and critical comments, as well as their participation in the audio-recordings. We thank the Service de Physiologie, Explorations Fonctionnelles et Médecine du Sport, Avicenne University Hospital, and the Clinical Research Unit of Avicenne University Hospital, for making the EEG recordings possible. LV acknowledges support from Fondation pour la Recherche Médicale (FRM Equipe grant), Inserm, CNRS, Collège de France and Fondation Bettencourt Schueller. JT acknowledges partial support from the National Science Foundation Division of Mathematical Sciences 1951369 and National Institute of Health R01GM152811. This clinical part of this work was supported by a grant from Contrat de Recherche Clinique 2021 (APHP211327). CP. was supported by a PhD grant from Ecole Normale Supérieure.

## Author contributions

Conceptualization: JT, CP, LV, BD; JT and CP carried out the conception and the design of the computational model; CP analyzed the simulations of the model; BD recruited the cohort of patients for EEG recordings; SNWT, ADL and BD performed EEG acquisition; CP analyzed the EEG data; CP wrote the manuscript. JT, LV, BD and CP have edited and corrected the manuscript.

