## Supplementary Information for "Deep Brain Stimulation restores information processing in parkinsonian cortical networks"

Charlotte Piette *et al*.

**This file includes:**

Figs. S1 to S7
Tables S1 to S6
Movies S1 to S2

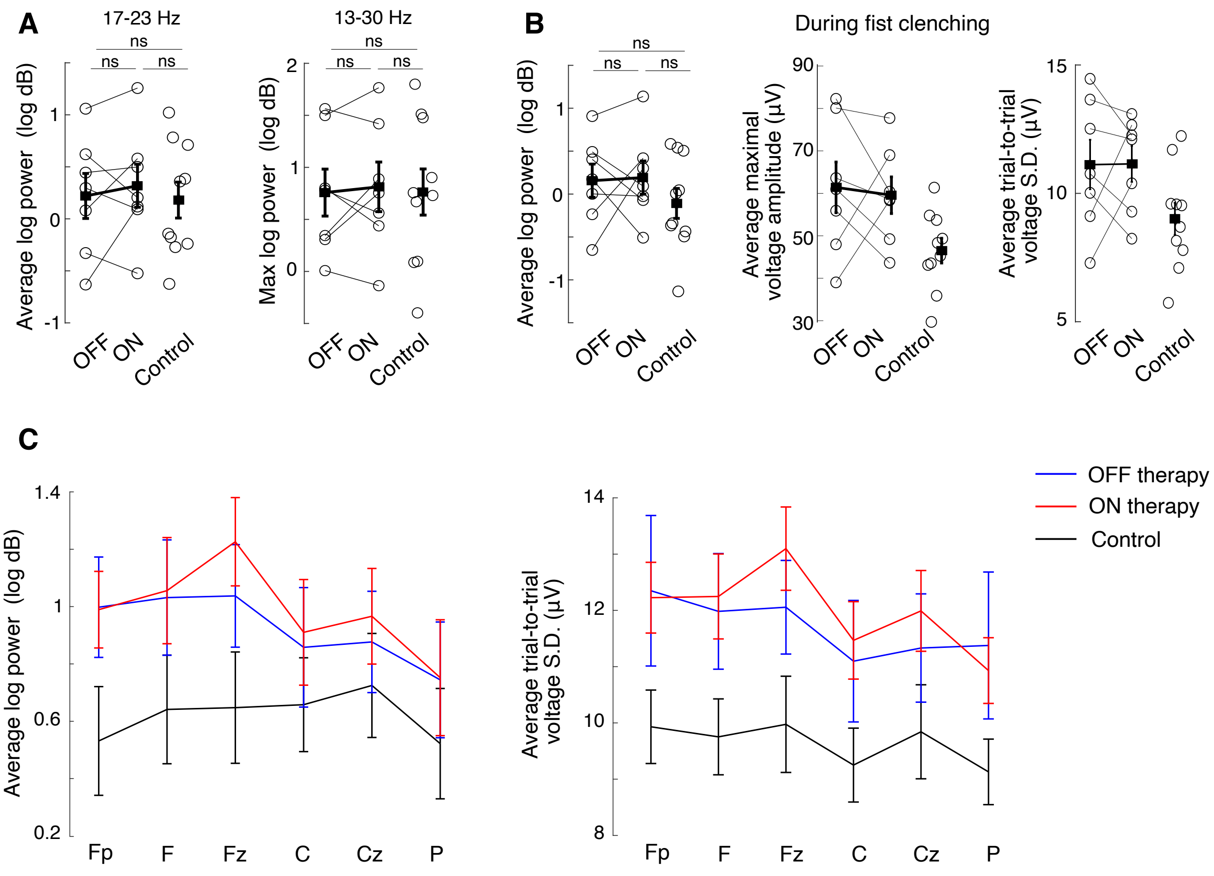


**Fig. S1. EEG features did not differ between DBS OFF and ON conditions.**

**A.** Alternative measures of beta band power across the entire finger tapping session, using the average log power over 17-23 Hz frequency range (left) or the maximal log power within the 13-30 Hz band (right) from the contralateral C electrode. No significant differences were observed between groups (paired Wilcoxon tests for OFF/ON condition: p=0.69 and p=0.69 respectively; independent Wilcoxon tests for OFF/Control: p=1 and p=0.96; for ON/Control: p=0.67 and p=0.96). **B.** EEG features measured from the contralateral C electrode during the initiation period of fist clenching show similar characteristics as for finger tapping. Left: Average beta-band power (13-30 Hz); Middle: Average maximal voltage amplitude; Right: Mean standard deviation of the voltage. Statistical tests: paired Wilcoxon tests for OFF/ON condition (p=0.94, p=0.69 and p=0.94); independent Wilcoxon tests for OFF/Control (p=0.42, p=0.55 and p=0.088) and for ON/Control (p=0.36, p=0.033 and p=0.043 respectively). **C.** The choice of analyzing the contralateral C electrode does not affect the observations previously made, as illustrated for the average beta-band power (left) and the mean standard deviation of the voltage (right).

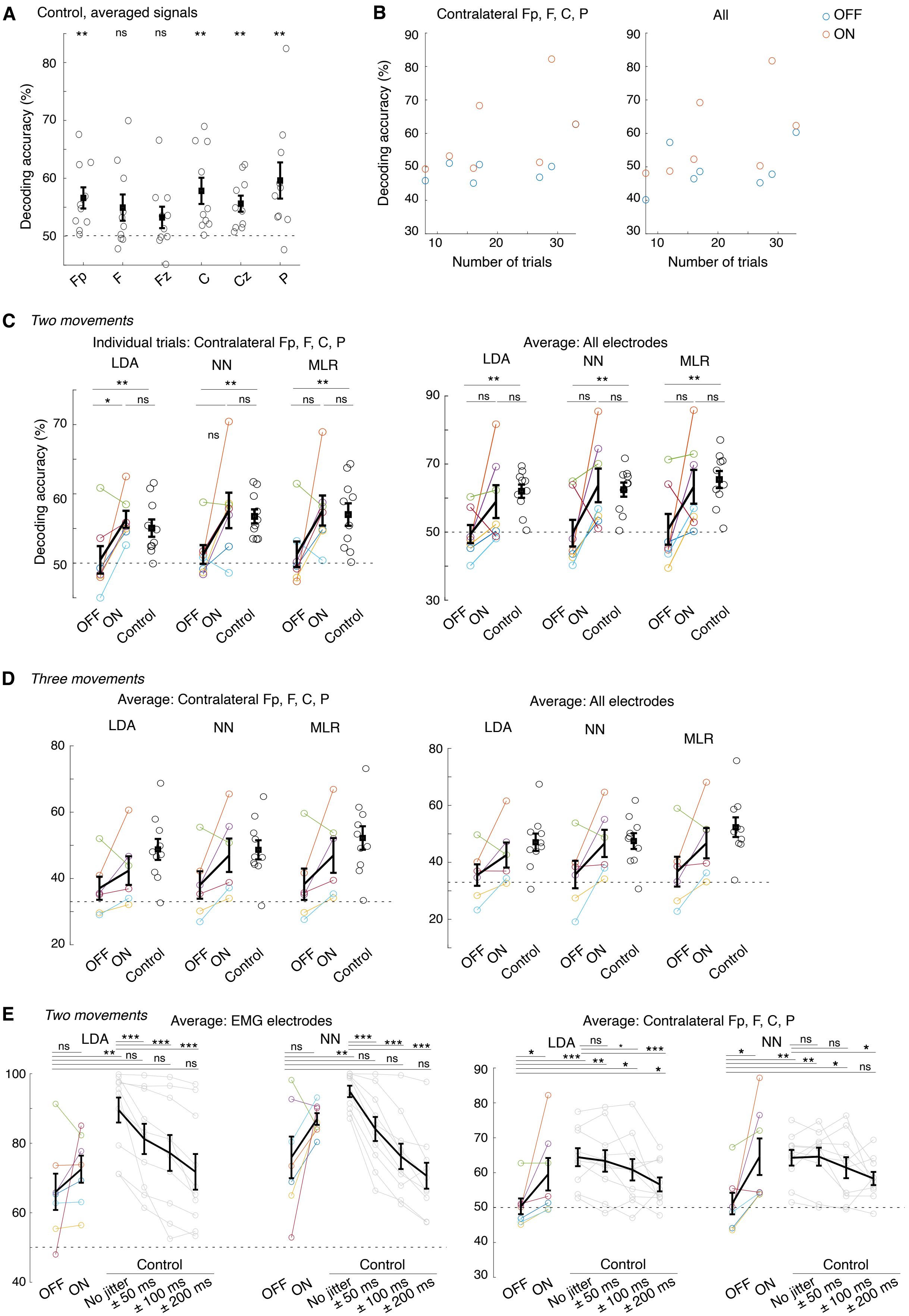
A
Control, averaged signals
Decoding accuracy (%)
Fp
F
Fz
C
Cz
P
** ns ns ** ** **
B
Contralateral Fp, F, C, P
All
OFF
ON
Decoding accuracy (%)
Number of trials
C
Two movements
Individual trials: Contralateral Fp, F, C, P
Average: All electrodes
LDA
NN
MLR
OFF
ON
Control
D
Three movements
Average: Contralateral Fp, F, C, P
Average: All electrodes
E
Two movements
Average: EMG electrodes
Average: Contralateral Fp, F, C, P
No jitter
± 50 ms
± 100 ms
± 200 ms

**Fig. S2. Altered decoding accuracy in movement identification from EEG signals in PD patients**

**A.** Decoding accuracy for control subjects (each dot corresponds to one subject) using linear discriminant analysis on individual electrode signals. One-sample Wilcoxon test (chance level: 50%, **Table S2**). **B.** For each patient, the number of templates for training the classifiers could differ (but remained similar between ON/OFF therapy), such that we ensured that the absence of correlation between decoding accuracy and number of training templates. Pearson's correlation coefficient r and *p*-value when trained on (left) contralateral Fp, F, C, P electrodes and (right) all electrodes: OFF: $r = 0.60$, $p = 0.15$ / ON: $r = 0.54$, $p = 0.22$; OFF: $r = 0.40$; $p = 0.37$; ON: $r = 0.55$; $p = 0.20$, respectively. **C.** Decoding accuracy across three classifiers between patients ON and OFF therapy (paired Wilcoxon test) and with control subjects (independent Wilcoxon test). See **Table S3** for detailed statistics. Individual signals from four electrodes contralateral to the movement side (left), and averaged signals from all electrodes (middle) or from the contralateral C electrode (right) were used. **D.** Same as panel C but for classifying three movements, and for averaged signals from four electrodes contralateral to the movement (left) or from all electrodes (right). **E.** Decoding accuracy across two classifiers between patients ON and OFF therapy (paired Wilcoxon test) and with control subjects (independent Wilcoxon test and non-parametric Friedman test). Control data were artificially jittered (jitter drawn from a uniform distribution bounded at ± 50, 100 or 200 milliseconds). See **Table S4** for detailed statistics. Individual signals from the three EMG electrodes ipsilateral to the movement side (left) or from four electrodes contralateral to the movement (right).

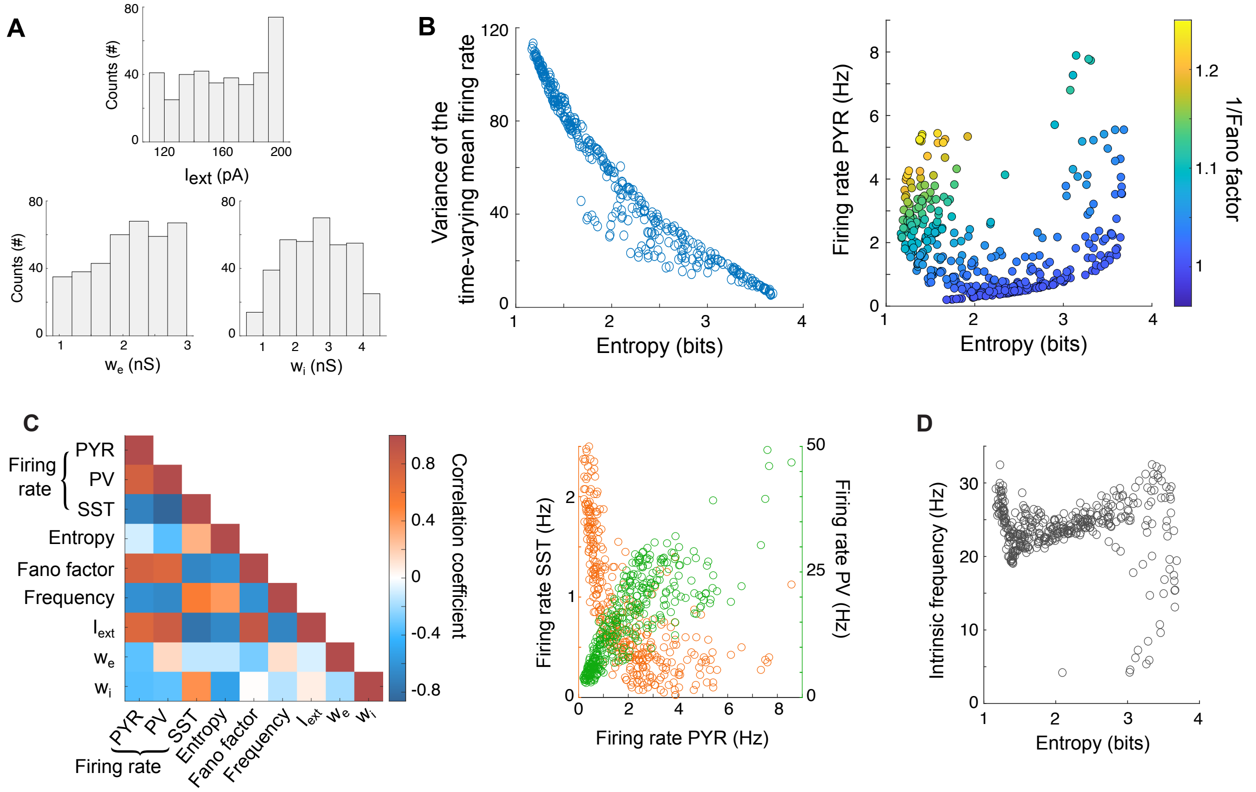


**Fig. S3. Analysis of the activity of network spiking models in the OFF DBS condition**
**A.** Distribution of the three variable parameters $I_{ext}$, $w_e$ and $w_i$, after exclusion of trials (*Methods*). **B.** Alternative measures of synchronization: (left) correlation between the average variance of pyramidal spike times and entropy; (right) 1/Fano factor (color-coded) mapped onto the entropy and firing rate of pyramidal cells. **C.** Left: Pearson's correlation coefficients between model parameters and network activity. Right: Correlation between the firing rates of pyramidal cells and PV neurons (Spearman's r = 0.87, $p < 0.001$), and SST interneurons (Spearman's r = -0.82, $p < 0.001$). **D**. Sharpening of the intrinsic frequency to beta frequency range as the level of synchronization increases (when the entropy decreases).

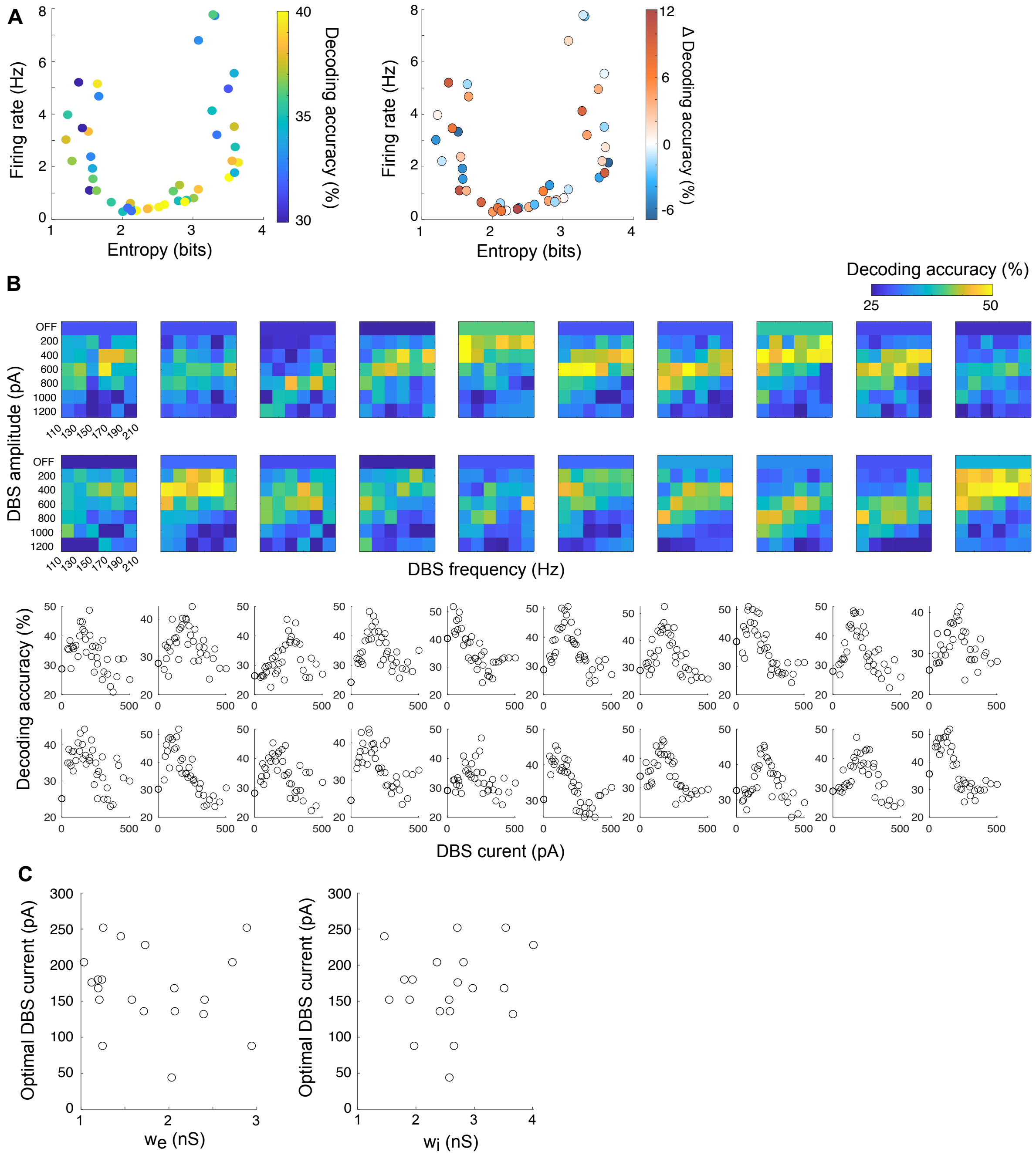


**Fig. S4. Finding optimal DBS parameters with naturalistic stimuli**

**A.** Left: Decoding accuracy when trained to recognize voices (3 voices, chance level: 33%) using multinomial logistic regression in the OFF DBS condition. Right: Changes in decoding accuracy under DBS (ON – OFF) when trained to recognize voices. Fixed DBS parameters (200 pA, 130 Hz) were used. **B.** Top: Decoding accuracy of the multinomial logistic regression when trained to recognize meaning (4 meanings, chance level: 25%) as a function of DBS frequency (x-axis) and amplitude (y-axis) for all models tested. Bottom: Projection of the decoding accuracy for all models on DBS current (x-axis), equal to DBS frequency x amplitude x pulse duration. **C**. Impact of network parameters $w_e$ (left) and $w_i$ (right) on the optimal DBS current (Pearson's correlation coefficient for $w_e$: r = -0.15, $p$ = 0.53; for $w_i$: r = 0.16, $p$ = 0.50).

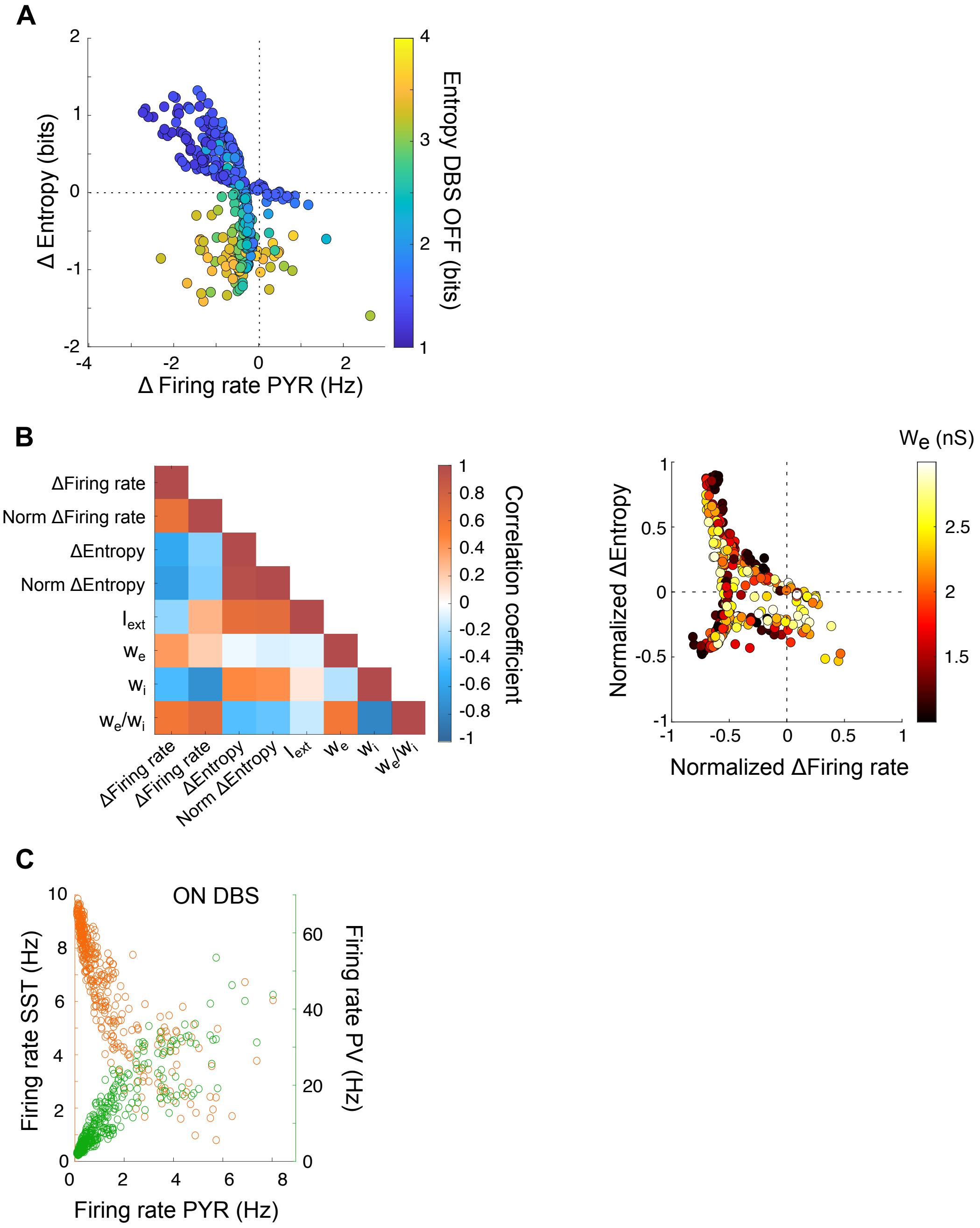


**Fig. S5. Impact of DBS on the activity of network spiking models**

**A.** Changes (absolute values) in entropy (y-axis) and firing rate (x-axis) between ON DBS and OFF DBS, as a function of the entropy OFF DBS (color-coded). **B.** Left: Correlation matrix between changes induced by DBS and network parameters (norm: normalized). High $w_i$ was correlated with a decrease in firing rate (r = -0.44, $p < 0.001$) and an increase in entropy (r = 0.46, $p < 0.001$). High $I_{ext}$ was especially correlated with an increase in entropy (r = 0.67, $p < 0.001$). Right: Impact of $w_e$ on DBS-mediated changes. **C.** Correlation between the firing rate of pyramidal cells and that of interneurons (SST in orange, PV in green) under high-frequency DBS. The same correlation structure was observed in the OFF DBS condition, despite the strong recruitment of SST interneurons.

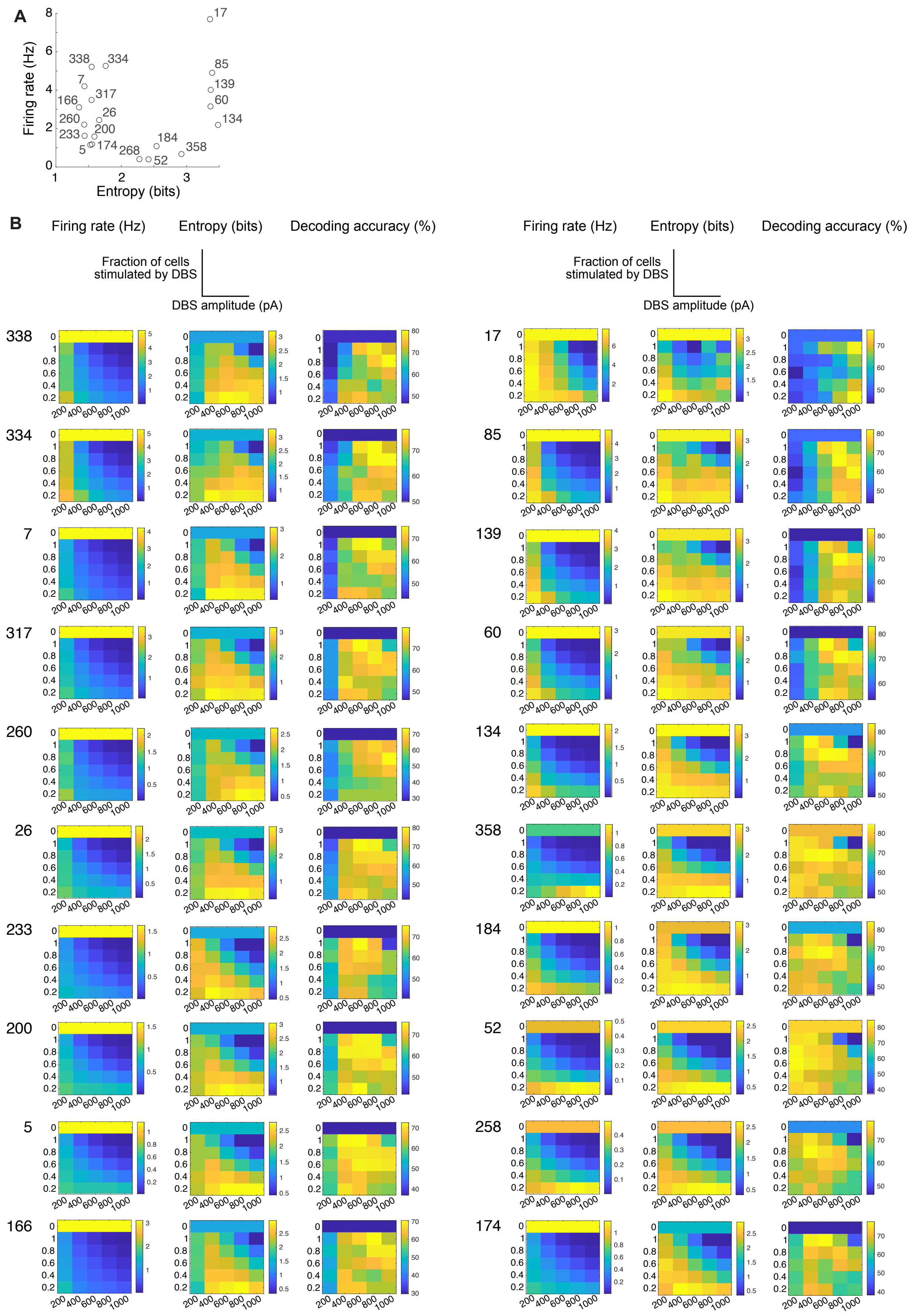
A
Firing rate (Hz)
Entropy (bits)
B
Firing rate (Hz)
Entropy (bits)
Decoding accuracy (%)
Fraction of cells stimulated by DBS
DBS amplitude (pA)
338
334
7
317
260
26
233
200
5
166
17
85
139
60
134
358
184
52
258
174

**Fig. S6. Impact of varying the proportion of neurons recruited under DBS**

**A.** Subset of network configuration in the firing rate/entropy diagram. Each network configuration is numbered. The 20 chosen configurations are scattered over the whole dynamic range observed from the set of 370 configurations. **B.** Evolution of the firing rate (Hz), entropy (bits) and decoding accuracy (LDA algorithm, %) as a function of the amplitude of DBS pulses (x-axis, DBS at 130 Hz) and the fraction of cells stimulated by DBS (y-axis) for each network configuration. Changes in firing rate, entropy and decoding accuracy under DBS are already markedly observed even when a small fraction of neurons is stimulated, without necessarily having to compensate with very high stimulation amplitude. For network configurations in which pyramidal cells show a high firing rate and DBS is less effective (eg. configurations 338, 334, 7, 17, 85, 139, 60), increasing DBS amplitude, even while maintaining a low fraction of stimulated cells, is particularly effective.

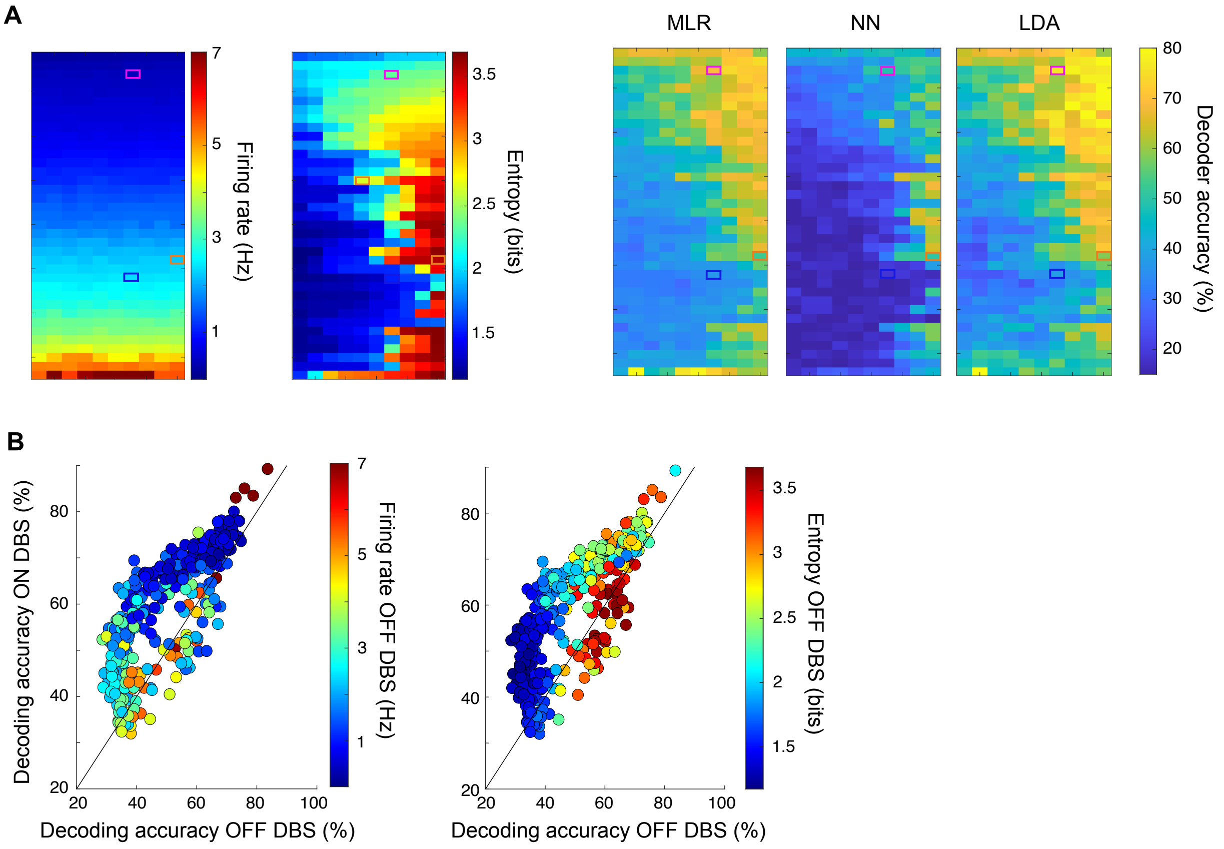


**Fig. S7. Decoding accuracy on simple stimuli as a function of the network's activity regime**

**A.** Network configurations ordered as a function of the firing rate and entropy (each square represents one network configuration). Decoding accuracy of the multinomial logistic regression (MLR), nearest-centroid (NN) and linear discriminant analysis (LDA) are compared using the same matrix arrangement. Overlaid colored rectangles refer to the three illustrative network profiles highlighted in **Fig. 2 & 4**. **B.** Comparison of MLR decoding accuracy OFF DBS (x-axis) and ON DBS (y-axis), with color-coded the firing rate OFF DBS (left) or the entropy OFF DBS (right).

| | Sex | Age | Equivalence DOPA (mg) | Start of disease (years) | Side for movement execution |
|---|---|---|---|---|---|
| DBS-implanted parkinsonian patients | 8 M, 2 F<br>Excluded: 3 M | 57.4 ± 2.8 (max: 71, min: 45) | 472 ± 98 | 7.1 ± 2.9 | 8 right<br>2 left |
| Control subjects | 8 M, 2 F | 57.7 ± 2.9 (max: 72, min: 45) | - | - | 9 right<br>1 left |

**Table S1. Characteristics of the cohort for EEG recordings**

| **Individual: 100 exemplars Average: 20 exemplars** | **Mean ± SEM (n=10 control)** | **Wilcoxon sign-rank test p-value** |
|---|---|---|
| **Figure 1E** | | |
| Individual, Contra C | 50.1 ± 0.5 | 0.5078 |
| Average, Contra C | 57.8 ± 2.3 | 0.002 |
| Individual, Contra Fp, F, C, P | 55.9 ± 2.4 | 0.0645 |
| Individual, Ipsi Fp, F, C, P | 52.2 ± 1.4 | 0.2324 |
| Average, Contra Fp, F, C, P | 60.8 ± 2.2 | 0.0059 |
| Average, All | 56.1 ± 2.3 | 0.0293 |
| Permutation, All | 58.5 ± 1.5 | 0.0059 |
| **Figure S2** | | |
| Permutation, Contra Fp | 56.6 ± 1.8 | 0.0020 |
| Permutation, Contra F | 54.9 ± 2.3 | 0.0804 |
| Permutation, Contra Fz | 53.2 ± 1.9 | 0.1602 |
| Permutation, Contra Cz | 55.6 ± 1.4 | 0.0020 |
| Permutation, Contra P | 59.6 ± 3.1 | 0.0039 |

**Table S2. Choice of electrode configuration for decoding movement**

| | Mean ± SEM<br><br>Per patient, training exemplars | **OFF therapy**<br><br>LDA<br>NN<br>MLR | **ON therapy**<br><br>LDA<br>NN<br>MLR | **Control**<br><br>LDA<br>NN<br>MLR | Paired Wilcoxon test OFF/ON<br>*p*-value | Wilcoxon test OFF/Control & ON/Control<br>*p*-value |
|---|---|---|---|---|---|---|
| 2 movements | Average, Contra Fp, F, C, P | 50.4 ± 2.3<br>51.1 ± 3.1<br>51.1 ± 4.0 | 59.5 ± 4.7<br>64.6 ± 5.2<br>63.8 ± 5.4 | 64.4 ± 2.6<br>64.3 ± 2.2<br>66.6 ± 2.8 | 0.0156<br>0.0312<br>0.0781 | <0.001 and 0.1932<br>0.0068 and 0.9623<br>0.0097 and 0.6009 |
| | Individual, Contra Fp, F, C, P | 50.4 ± 2.0<br>51.1 ± 1.4<br>51.3 ± 1.8 | 56.3 ± 1.2<br>57.6 ± 2.6<br>57.6 ± 2.2 | 55.1 ± 1.3<br>56.7 ± 1.0<br>57.0 ± 1.6 | 0.0469<br>0.0781<br>0.0781 | 0.0431 and 0.3148<br>0.0046 and 0.8868<br>0.0025 and 0.9623 |
| | Average, All | 49.4 ± 2.7<br>49.7 ± 3.9<br>50.8 ± 4.5 | 59.0 ± 4.8<br>63.8 ± 4.9<br>63.3 ± 5.0 | 62.0 ± 1.9<br>62.5 ± 2.1<br>65.5 ± 2.5 | 0.1562<br>0.1094<br>0.0781 | 0.002 and 0.3638<br>0.0136 and 0.9623<br>0.033 and 0.6009 |
| | Average, Contra C | 50.4 ± 2.6<br>51.1 ± 3.5<br>51.0 ± 3.3 | 56.5 ± 3.4<br>60.8 ± 4.4<br>59.2 ± 3.9 | 64.4 ± 2.6<br>64.3 ± 2.2<br>66.6 ± 2.8 | 0.2188<br>0.2188<br>0.2969 | 0.0031 and 0.0553<br>0.0185 and 0.4173<br>0.0046 and 0.1932 |
| 3 movements* | Average, Contra Fp, F, C, P | 37.1 ± 3.7<br>38.0 ± 4.8<br>38.2 ± 5.2 | 42.4 ± 4.3<br>47.0 ± 5.0<br>46.9 ± 5.2 | 48.8 ± 3.2<br>48.6 ± 2.9<br>52.2 ± 3.5 | 0.2188<br>0.1562<br>0.1562 | 0.0420 and 0.2198<br>0.0420 and 0.7925<br>0.0420 and 0.4923 |
| | Average, All | 35.5 ± 3.7<br>35.7 ± 4.8<br>36.7 ± 5.2 | 42.6 ± 4.4<br>46.6 ± 4.8<br>46.7 ± 5.3 | 47.0 ± 3.0<br>47.5 ± 2.8<br>52.3 ± 3.4 | 0.2188<br>0.0938<br>0.1562 | 0.0312 and 0.3132<br>0.0420 and 0.7128<br>0.0312 and 0.4278 |

*For two movements: n=7 OFF/ON therapy; for three movements: n=6 OFF/ON therapy. In control patients, n=10.

**Table S3. Summary of movement decoding accuracy from EEG signals**

| Mean ± SEM | OFF therapy | ON therapy | Control<br>LDA<br>NN | | | | Paired Wilcoxon test OFF/ON $p$-value | Non-parametric Friedman test (Control) | Wilcoxon test OFF/Control $p$-value | | | |
|---|---|---|---|---|---|---|---|---|---|---|---|---|
| Per patient, training exemplars | LDA<br>NN | LDA<br>NN | No jitter | ± 50 ms | ± 100 ms | ± 200 ms | | | | | | |
| Average, EMG electrodes | 66.0 ± 5.2<br>75.9 ± 6.0 | 72.5 ± 3.9<br>87.0 ± 1.7 | 89.6 ± 3.6<br>94.9 ± 1.6 | 81.2 ± 4.3<br>84.1 ± 3.5 | 77.2 ± 5.2<br>76.2 ± 3.6 | 71.8 ± 5.1<br>70.6 ± 3.7 | 0.156<br>0.219 | $\chi^2 = 27.7$<br>$p<0.001$<br>$\chi^2 = 27.7$<br>$p<0.001$ | 0.00308<br>0.00967 | 0.0553<br>0.315 | 0.193<br>0.475 | 0.601<br>0.219 |
| Average, Contra Fp, F, C, P | 50.4 ± 2.3<br>51.1 ± 3.1 | 59.5 ± 4.7<br>64.6 ± 5.2 | 64.4 ± 2.6<br>64.3 ± 2.2 | 63.4 ± 3.1<br>64.6 ± 2.6 | 60.8 ± 3.1<br>61.4 ± 3.1 | 56.6 ± 2.0<br>58.3 ± 1.9 | 0.0156<br>0.0313 | $\chi^2 = 10.2$<br>$p=0.017$<br>$\chi^2 = 6.96$<br>$p=0.073$ | <0.001<br>0.00679 | 0.00967<br>0.00967 | 0.0136<br>0.0330 | 0.0136<br>0.0553 |

**Table S4. Comparison of movement decoding accuracy from EMG and EEG signals in presence of jitter**

| | Pyramidal cells | PV cells | SST cells |
|---|---|---|---|
| $g_{leak}$ (nS) | 6 | 5 | 5 |
| $C$ (pF) | 180 | 80 | 80 |
| $V_{thres}$ (mV) | -49 | -52 | -53 |
| $\Delta_{thres}$ (mV) | 1 | 1 | 5 |
| $a$ (nS) | 4 | 0 | 4 |
| $b$ (pA) | 100 | 0 | 90 |
| $\tau_w$ (ms) | 100 | 15 | 40 |
| σ | 5 | 5 | 5 |
| $I_{ext}$(pA) | variable | 50 | 25 |

**Table S5. Intrinsic parameters of the model for each neuronal population**

| ***p, w*** | Pyramidal cells | PV cells | SST cells |
|---|---|---|---|
| Pyramidal cells | $p$ = 0.5, $w$ =0.5 nS | $p$ = 0.4, variable $w_e$ | - |
| PV cells | $p$ =0.4, variable $w_i$ | $p$ = 0.6, $w$= 2.5 nS | $p$ = 0.1, w = 1.6 nS |
| SST cells | $p$ = 0.4, $w$ = 2.2 nS | $p$ = 0.3, $w$ = 2.4 nS | $p$= 0.1, w = 1.6 nS |

**Table S6. Connectivity parameters between neuronal populations**

**Movie S1.**
Example of movement execution from a control patient

**Movie S2.**
Example of movement execution from a parkinsonian patient in OFF and ON therapy
